# Comparative effectiveness of ARB and ACEi for cardiovascular outcomes and risk of angioedema among different ethnic groups in England: an analysis in the UK Clinical Practice Research Datalink with emulation of a reference trial (ONTARGET)

**DOI:** 10.1101/2024.01.17.24301397

**Authors:** Paris J Baptiste, Angel YS Wong, Anna Schultze, Catherine M Clase, Clémence Leyrat, Elizabeth Williamson, Emma Powell, Johannes FE Mann, Marianne Cunnington, Koon Teo, Shrikant I Bangdiwala, Peggy Gao, Kevin Wing, Laurie Tomlinson

## Abstract

**Objective:** To study the comparative effectiveness of angiotensin receptor blockers (ARB) and angiotensin-converting enzyme inhibitors (ACEi) in ethnic minority groups in the UK.

**Design:** Observational cohort study using a reference trial emulation approach benchmarked against the ONTARGET trial.

**Setting:** UK Clinical Practice Research Datalink Aurum data from 01/01/2001-31/07/2019.

**Participants:** Black, South Asian, or White patients treated with ARB or ACEi who met the ONTARGET trial criteria.

**Main outcome measures:** The primary composite outcome was: cardiovascular-related death, myocardial infarction, stroke, or hospitalisation for heart failure with individual components studied as secondary outcomes. Angioedema was a safety endpoint. We assessed outcomes using a propensity-score—weighted Cox proportional hazards model for ARB vs ACEi with heterogeneity by ethnicity assessed on the relative and absolute scale.

**Results:** 17,593 Black, 30,805 South Asian, and 524,623 White patients were included. We benchmarked results against ONTARGET comparing ARB with ACEi for the primary outcome (hazard ratio [HR] 0.96, 95% CI: 0.95 to 0.98) and found no evidence of treatment effect heterogeneity (*P_int_*=0.422). Results were consistent for most secondary outcomes. However, for cardiovascular-related death, there was strong evidence of heterogeneity (*P_int_*=0.002), with ARB associated with more events in Black individuals and with fewer events in White individuals compared to ACEi, and no differences in South Asian individuals. For angioedema, HR 0.56 (95% CI: 0.46 to 0.67) for ARB vs ACEi (*P_int_*=0.306). Absolute risks were higher in Black individuals, for ARB vs ACEi number-needed-to-treat was 204 in Black individuals compared with 2000 in South Asian individuals and 1667 in White individuals (*P_int_*=0.023).

**Conclusions:** These results demonstrate variation in drug effects of ACEi and ARB by ethnicity and suggest the potential for adverse consequences from current UK guideline recommendations for ARB in preference to ACEi for Black individuals.

**What is already known on this topic**

- Current UK National Institute for Health and Care Excellence (NICE) recommendations for treatment of hypertension includes ethnicity (Black vs non-Black) as a determinant of treatment choice and recommend an ARB in preference to an ACEi in Black patients based on the risk of angioedema in this group
- Despite being at increased risk of hypertension and cardiovascular disease, little is known about comparative treatment effectiveness and risk of ARB and ACEi among South Asian patients in the UK
- Reference trial emulation (considering study design and benchmarking against an existing randomised trial), followed by analysis of effects in trial-underrepresented groups can add confidence to findings of observational research and bridge gaps in evidence

**What this study adds**

- In this propensity-score—weighted cohort study of self-reported Black, South Asian and White patients at high-risk of cardiovascular disease in the UK, ARB were as effective as ACEi at preventing most cardiovascular outcomes. However, for cardiovascular death, for Black patients the number-needed-to-harm (NNH) was 93 (95% CI: 49 to 1000) for ARB compared to ACEi. The opposite effect was observed in White patients: number-needed-to-treat (NNT) was 115 (95% CI: 91 to 159) for ARB vs ACEi to prevent one additional cardiovascular death. No difference between the two drugs was observed among South Asian patients.
- Relative risks of angioedema, ARB vs ACEi were similar across all ethnic groups but because of the increased incidence in Black patients, there was a marked difference in the NNT. For ARB compared to ACEi use, NNT 204 (95% CI: 127 to 556) in Black patients and NNT 1667 (95% CI: 1111 to 3333) in White patients, to prevent one additional angioedema outcome. There was no difference in angioedema risk between the drugs in South Asian patients.
- South Asian and White patients starting ACEi experienced a greater fall in blood pressure than those starting ARB. There was no difference in fall in blood pressure between the drugs for Black patients.

## Introduction

Hypertension is associated with increased cardiovascular risk.[1, 2] Individuals of African and Caribbean descent (subsequently referred to as ‘Black’), and of South Asian descent are disproportionately affected by hypertension in comparison with White individuals.[3] The extent to which these differences are related to genetics, differences in socio-economic status[4] or factors such as differential access to healthcare[5–7] is uncertain.[8–10] Incidence and mortality from hypertension and stroke is increased among Black and South Asian ethnic groups and occurs at a younger age.[11–13] In the UK, patients with hypertension are treated based on the National Institute for Health and Care Excellence (NICE) hypertension guidelines.[14] In contrast to other international guidelines, NICE includes ethnicity as a determinant of treatment choice, although the evidence underpinning this is uncertain.[15, 16] Despite this there are known variations to the renin-angiotensin system among people of different ethnicities, making variation of drug effects biological plausible.[17] Among patients with hypertension and type 2 diabetes, an angiotensin-converting enzyme inhibitor (ACEi) or angiotensin receptor blocker (ARB) is recommended as first-line treatment. Among patients with hypertension but without type 2 diabetes, an ACEi or ARB is recommended as initial treatment if the patient is aged <55 years and not Black, with calcium channel blocker being recommended to those aged ≥55 years or those who are Black of any age. In 2011 guidelines were updated to recommend an ARB in preference to an ACEi for Black people.[18] The cited evidence was the US ALLHAT trial of 42,418 patients in which over a third of participants were Black.[19] This trial found that over half of people who developed angioedema were Black[20] and that the incidence of angioedema was higher among Black ACEi users compared with users of other antihypertensive drugs (not including ARB), which was not the case in non-Black participants. However, the absolute incidence was low with only 53 events during a follow-up of 4.9 years.

Trials demonstrating the comparative effectiveness of ARB and ACEi which inform clinical guidance have provided limited data on the effects in Black or South Asian people.[21–23] The global ONTARGET trial, which demonstrated non-inferiority of telmisartan compared with ramipril in participants at high-risk of cardiovascular events, did not include subgroup analyses by ethnicity and only 2% of included participants were Black (reported in the trial as Black African). This is consistent with the majority of clinical trials, which have historically reported limited subgroup analyses by race or ethnicity.[24] This practice can lead to extrapolation of trial results to ethnic minority populations without robust evidence. Initiatives have been put in place to improve diversity in clinical trials.[25, 26] A further approach to bridge this gap in evidence is to explore drug effects in diverse populations using observational studies in routine-care data.

Reference trial emulation in comparative effectiveness research involves use of an existing named RCT to (1) inform observational study design and (2) benchmark results against, providing confidence in validity of the selected observational methods and data. Analysis can then be extended to study treatment effects within underrepresented subgroups, in this case ethnic minority groups, with the large sample sizes and diverse population characteristic of UK routinely collected data maximising power for these analyses.[27–30]

We sought to determine whether ARB and ACEi were equally effective for reducing cardiovascular and renal outcomes, and to quantify the risk of angioedema, among White, Black and South Asian people in the UK Clinical Practice Research Datalink Aurum, applying reference trial emulation to observational data to provide confidence in the validity of results.

## Methods

### The reference RCT (ONTARGET)

#### ONTARGET methods

The primary objective of the ONTARGET trial was to determine if telmisartan (ARB) was non-inferior to ramipril (ACEi) for reduction of cardiovascular events among patients aged ≥55 years with vascular disease or high-risk diabetes, but without heart failure.[23] The primary outcome of the trial was a composite of cardiovascular-related death, myocardial infarction (MI), stroke or hospitalisation for heart failure.

#### ONTARGET results

Among the participants included in the trial 1.2% were of South Asian and 2% were of Black ethnicity. Just over 8500 patients were enrolled into each of the treatment arms of ramipril or telmisartan. The primary outcome occurred in 1412 (16.5%) patients in the ramipril group and 1423 (16.7%) patients in the telmisartan group and showed HR 1.01 (95% CI: 0.94 to 1.09) for telmisartan 80 mg daily vs ramipril 10 mg daily.[23]

### Data sources

CPRD Aurum was used, rather than GOLD, to increase sample size and power.[16] As of June 2021, CPRD Aurum includes primary care records for research-acceptable patients from England (registered at currently contributing practices, excluding deceased patients) representative of around 20% of the UK population.[31] This was linked to hospitalisation and mortality data from HES and the Office for National Statistics (ONS).

### Outcomes

Comparisons were made between ARB vs ACEi and outcomes replicated those studied in ONTARGET.

- Primary outcome: composite of cardiovascular death, myocardial infarction (MI), stroke, or hospital admission for congestive heart failure
- Secondary cardiovascular outcomes:

- Main secondary outcome: composite of cardiovascular death, MI, or stroke
- Individual components of primary outcome
- Secondary kidney outcomes:

- Loss of glomerular filtration rate (GFR) or development of end-stage kidney disease (ESKD) (defined as: ≥50% reduction in estimated GFR (eGFR), start of kidney replacement therapy (KRT) or development of eGFR <15ml/min/1.73m^2^)
- Development of ESKD (defined as: start of KRT or development of eGFR <15ml/min/1.73m^2^)
- Doubling of serum creatinine

GFR was calculated using the CKD-Epi equation 2009 without reference to ethnicity.[32]

- Other outcomes:

- Death from non-cardiovascular causes
- All-cause mortality
- Safety outcome: angioedema

### Emulation of the reference RCT

Our methods for emulation of the reference trial (ONTARGET) in CPRD Aurum are summarised below. Full details can be found in a previously published protocol and summarised in Figure S1 of the supplementary material.[33] Supplementary Table S1 details protocol components from: ONTARGET and the emulation in CPRD Aurum.

#### Eligibility criteria and treatment strategies

##### Step 1: Create exposed periods

We selected Black, South Asian and White patients ever prescribed an ARB and/or ACEi between 2001-01-01 to 2019-07-31 in CPRD Aurum, with ethnicity defined using both CPRD and HES to improve completeness.[34] Ethnicity was self-reported; those with missing ethnicity were excluded. Courses of ARB or ACEi therapy were denoted as exposed periods, a new exposed period was considered when a treatment gap of more than 90 days occurred. Therefore, a patient could contribute multiple eligible exposed periods, as in a trial a patient could meet the trial criteria on more than one occasion.

##### Step 2: Create trial-eligible periods

We applied the ONTARGET trial criteria to the start of each exposed period to select high-risk patients aged ≥55 years with a previous cardiovascular diagnosis, or diabetes with complications. Table S10-S11 of the supplementary material shows how trial criteria were interpreted in CPRD.

#### Statistical analysis

##### Step 3: Balance across exposure groups

From the trial-eligible periods defined in step 2, we selected one random eligible period per patient and developed a propensity score model for the probability of receiving an ACEi within each ethnicity group strata (White, Black and South Asian). Variables considered in the propensity score model included demographics, medication and clinical history, and time-related variables to account for bias introduced in treatment switchers (supplementary Table S2). Propensity scores were trimmed at the 1^st^ percentile in the ACEi group and the 99^th^ percentile in the ARB group to avoid extreme weights and violation of the positivity assumption. Treatment groups were weighted by propensity score to obtain balance of baseline characteristics.

##### Benchmarking results

We explored the replicability of the ONTARGET trial findings in our trial-eligible cohort by estimating a hazard ratio (HR) using the Cox-proportional hazards model, weighted by propensity score, for the primary composite trial outcome of cardiovascular death, MI, stroke or hospitalisation for heart failure and for components of this outcome separately, in addition to the main secondary outcome. Patients were followed from the start of the trial-eligible period until the first of: outcome, death, transfer out of practice, last collection date or 5.5 years. We confirmed similarity to the trial if our results for the primary composite outcome met a pre-specified validation criteria of 1). HR between 0.92 and 1.13 and 2). 95% CI for the HR contained 1.[33] No criteria were pre-specified to confirm replicability of the secondary outcomes other than investigators’ judgement that results were consistent with the RCT. Only outcomes studied in ONTARGET were assessed in this benchmarking step. These methods mirrored those that were implemented in a previous analysis using CPRD GOLD.[33, 35] Data were analysed using Stata/MP version 17.0.

### Extending analysis to the underrepresented ethnic groups

After benchmarking results against the reference RCT, analysis was extended to the underrepresented ethnic groups.

#### Eligibility criteria and treatment strategies

We used the same CPRD Aurum cohort prepared for the reference trial emulation (i.e. data sources, outcomes, eligibility and treatment strategies as described above).

#### Statistical analysis

Benchmarking of results by ethnic group was not performed (as the extended analysis was specifically to perform analysis by ethnic group that was not performed in the trial). We explored treatment effect heterogeneity on the multiplicative scale by ethnic group using a Wald test for an interaction between treatment and ethnic group in the weighted Cox-proportional hazard model, showing relative differences between groups.

Unlike other adverse events drug-related angioedema can occur years after treatment.[36] Therefore, we examined the risk of developing angioedema over the total follow-up period of 5.5 years. After identifying ethnic differences in the frequency of angioedema, we decided to report also propensity-score—weighted incidence rates using a Poisson regression model, examining the absolute differences between groups, so that we could interpret results in terms of numbers-needed-to-treat (NNT) and numbers-needed-to-harm (NNH).

We reported changes in blood pressure from baseline by treatment and ethnicity. These were assessed at (or closest to) 6 weeks, 6 months, 12 months, 18 months, 2 years, 3 years and 4 years after the start of the trial-eligible period.

### Sensitivity analyses

We confirmed our findings by comparing the main intention-to-treat analysis results from our CPRD cohort with results obtained under an on-treatment analysis. This involved additionally censoring patients if they ended treatment, switched, or became a dual user of ACEi/ARB. End of treatment was defined as the end of eligible period, i.e., when a treatment gap of >90 days occurred. Censor date was defined as date of last dose of study drug + 60 days.

To assess the bias introduced by including variables with missing values in our propensity-score model, we repeated analyses after multiple imputation of baseline blood pressure and creatinine, for which we judged it reasonable to assume missing at random. Other variables originally included in the model had <10% missing data so complete cases were included.[37, 38] Values were imputed for baseline variables only, therefore this analysis was assessed for non-kidney related outcomes only.

We assessed the impact that the 2011 guideline update (recommending ARB to Black patients in preference over an ACEi)[18] might have had on results by restricting the cohort to trial-eligible periods prior to 2011 for the primary cardiovascular outcome when extending analysis to underrepresented ethnic groups.

### Patient and public involvement

No patients were involved in setting the research question or the outcome measures, nor were they involved in developing plans for design or implementation of the study. No patients were asked to advise on interpretation or writing up of results.

## Results

### Baseline characteristics

In total 573,021 patients were included in the analysis, of whom 74% were prescribed an ACEi. Among the cohort, 17,593 were Black, 30,805 were South Asian and 524,623 were White (Figure 1, Table 1). ACEi continued to be prescribed more than ARB between 2001-2018 for all ethnic groups. After the 2011 treatment recommendation update,[18] a small increase in ARB prescriptions were observed among Black individuals. Prescribing patterns were similar among Black and South Asian individuals (supplementary Figure S2). South Asian individuals were youngest of the three ethnic groups studied (supplementary Table S4-S6). A greater proportion of Black and South Asian individuals met the trial inclusion criteria because of high-risk diabetes, in comparison with White individuals, whose main reason for inclusion was coronary artery disease. Black and South Asian individuals were less likely to smoke or drink, had more appointments in primary care, and Black individuals were the most deprived (supplementary Tables S4-S6).

**Figure 1.**
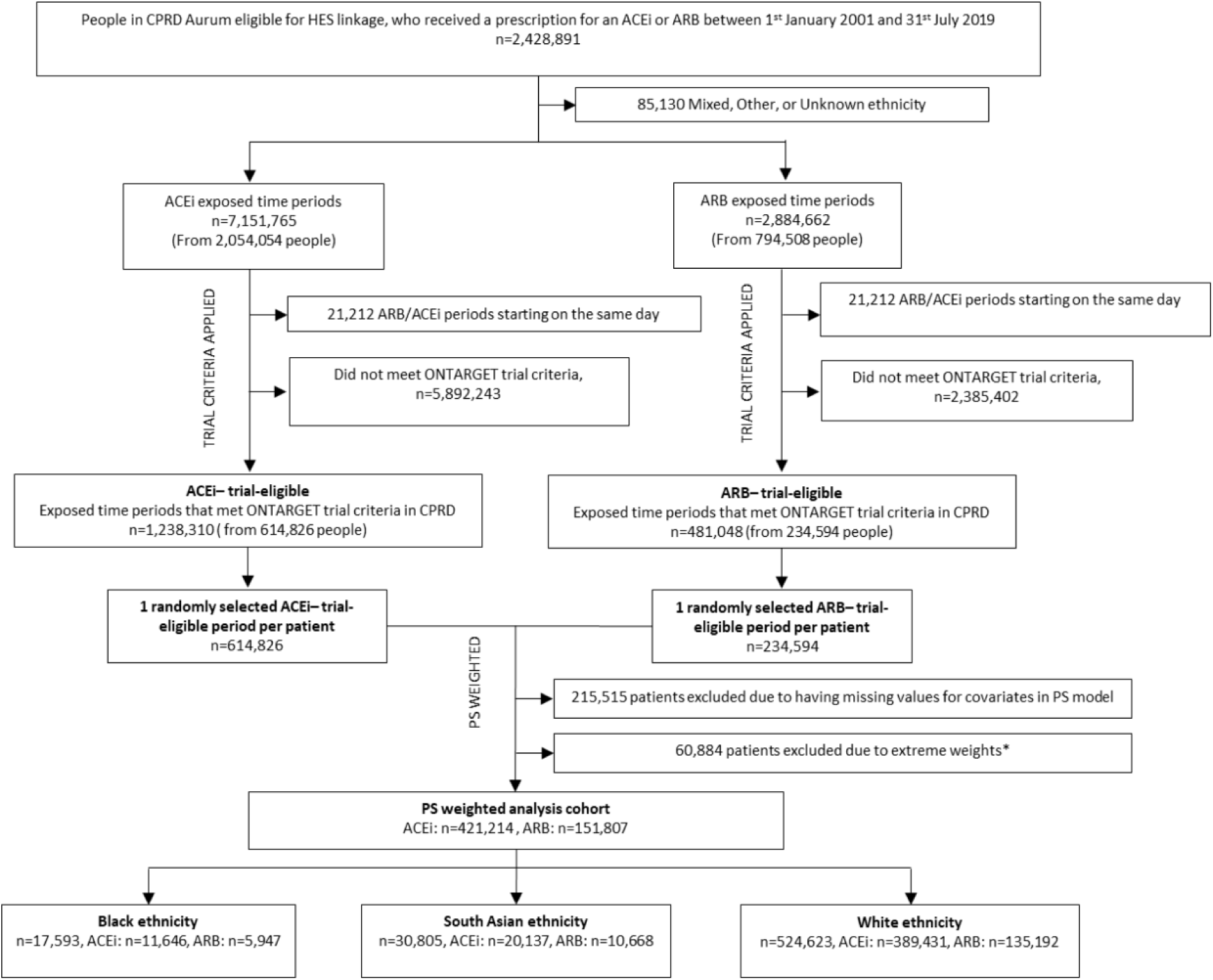
Study diagram for people included in propensity-score—weighted analysis cohort using CPRD Aurum. Notes: ARB= angiotensin receptor blocker; ACEi= angiotensin-converting enzyme inhibitor; PS= propensity score. *propensity scores were trimmed at the 1^st^ percentile in the ACEi group and at the 99^th^ percentile in the ARB group to avoid extreme weights.

**Table 1.** Baseline characteristics of trial-eligible patients after applying trial criteria included in propensity-score—weighted reference trial emulation analysis compared to ONTARGET.

| Characteristic | ARB<br>N=151,807 | ACEi<br>N=421,214 | ONTARGET<br>N=25,620 |
| --- | --- | --- | --- |
| Age (year) – mean (SD) | 71.3 (9.2) | 70.9 (9.4) | 66.4 (7.2) |
| Systolic BP (mmHg) – mean (SD) | 144.0 (20.2) | 143.6 (20.2) | 141.8 (17.4) |
| Diastolic BP (mmHg) – mean (SD) | 78.8 (10.9) | 79.1 (11.0) | 82.1 (10.4) |
| Body-mass index – mean (SD) | 29.2 (5.8) | 28.8 (5.8) | 28.2 (4.7) |

| **Characteristic** | **ARB**<br>N=151,807 | **ACEi**<br>N=421,214 | **ONTARGET**<br>N=25,620 |
| --- | --- | --- | --- |
| **Creatinine (μmol/l) – mean (SD)** | 94.1 (29.5) | 92.9 (27.3) | 94.2 (24.4) |
| **Female sex – no. (%)** | 82849 (54.6) | 202511 (48.1) | 6831 (26.7) |
| **Ethnic group – no. (%)** |  |  |  |
| Black | 5947 (3.9) | 11646 (2.8) | 511 (2.0) |
| Other | 0 | 0 | 5973 (23.3) |
| South Asian | 10668 (7.0) | 20137 (4.8) | 303 (1.2) |
| Unknown | 0 | 0 | 125 (<0.1) |
| White | 135192 (89.1) | 389431 (92.5) | 18708 (73.0) |
| **Clinical history – no. (%)** |  |  |  |
| CAD<sup>a</sup> | 105969 (69.8) | 298697 (70.9) | 19102 (74.6) |
| Cerebrovascular disease<sup>b</sup> | 16071 (10.6) | 44821 (10.6) | 5342 (20.9) |
| PAD<sup>c</sup> | 14158 (9.3) | 38958 (9.3) | 3468 (13.5) |
| Diabetes | 93709 (61.7) | 250030 (59.4) | 9612 (37.5) |
| High-risk diabetes<sup>d</sup> | 76297 (50.3) | 197471 (46.9) | 7151 (27.9) |
| **Smoking status – no. (%)** |  |  |  |
| Non-smoker | 44137 (29.1) | 113688 (27.0) | 9088 (35.5) |
| Current smoker | 35111 (23.1) | 111998 (26.6) | 3225 (12.6) |
| Past smoker | 72559 (47.8) | 195528 (46.4) | 13276 (51.8) |
| Unknown | 0 | 0 | 31 (0.1) |
| **Alcohol status – no. (%)** |  |  |  |
| Yes | 92697 (61.1) | 261221 (62.0) | 10345 (40.4) |
| No | 45266 (29.8) | 119770 (28.4) |  |
| Unknown | 13844 (9.1) | 40223 (9.6) | 14 (<0.1) |
| **Medication<sup>e</sup> – no. (%)** |  |  |  |
| Alpha-blocker | 17089 (11.3) | 38166 (9.1) | 1095 (4.3) |
| Oral anticoagulant agent | 13055 (8.6) | 34986 (8.3) | 1939 (7.6) |
| Antiplatelet agent | 13365 (8.8) | 40980 (9.7) | 2824 (11.0) |
| Aspirin | 50393 (33.2) | 148837 (35.3) | 19403 (75.7) |
| Beta-blocker | 47734 (31.4) | 135829 (32.3) | 14583 (56.9) |
| Calcium-channel blocker | 52535 (34.6) | 135915 (32.3) | 8472 (33.1) |
| Digoxin | 5430 (3.6) | 16976 (4.0) | 865 (3.4) |
| Diuretics | 63949 (42.1) | 163355 (38.8) | 7164 (28.0) |
| Diabetic treatment | 38321 (25.2) | 101647 (24.1) | 8056 (31.4) |
| Nitrates | 14102 (9.3) | 43919 (10.4) | 7523 (29.4) |
| Statins | 79774 (52.6) | 225469 (53.5) | 15783 (61.6) |

| Characteristic | ARB<br>N=151,807 | ACEi<br>N=421,214 | ONTARGET<br>N=25,620 |
| --- | --- | --- | --- |
| N= number of patients; SD=standard deviation; no. (%)=number (percent); BP= blood pressure; CAD=coronary artery disease; PAD=peripheral artery disease; CKD=chronic kidney disease (eGFR<60ml/min/1.73m<sup>2</sup>)<br>One third of ONTARGET participants received both ramipril plus telmisartan.<br><sup>a</sup> Includes diagnosis of: MI at least 2 days prior, angina at least 30 days prior, angioplasty at least 30 days prior, CABG at least 4 years prior<br><sup>b</sup> Includes diagnosis of: stroke/TIA<br><sup>c</sup> Includes diagnosis of: limb bypass surgery, limb/foot amputation, intermittent claudication<br><sup>d</sup> Includes DM with: retinopathy, neuropathy, chronic kidney disease, proteinuria or other complication<br><sup>e</sup> Within 3 months prior to eligible start date. Antiplatelet agent= clopidogrel/ticlopidine<br>Black ethnic group presented for ONTARGET includes ‘Black African’ and White ethnic group presented for ONTARGET includes ‘European/Caucasian’ as described in the trial protocol. South Asian ethnic group presented for ONTARGET includes Indian, Sri Lanka, Pakistan, Bangladesh, Afghanistan, and Nepal. ONTARGET additionally included ‘Colored African’ ethnicity in the CRF which we re-categorised to unknown in this table. |  |  |  |

### Emulation of the reference RCT

Baseline characteristics and standardised differences after weighting are shown in Tables S3-S6 of the supplementary material.

In the whole study population, the primary composite outcome occurred in 27,327 (18.0%) in the ARB group and in 80,624 (19.1%) in the ACEi group, representing event rates of 25% and 26% per 5.5 person-years in the ARB and ACEi exposure groups, respectively over a median follow-up of 5.2 years.

#### Benchmarking CPRD Aurum results against the ONTARGET trial results

The estimated HR was 0.96 (95% CI: 0.95 to 0.98) for the comparison of ARB vs ACEi for the primary composite outcome (supplementary Table S7), this met the first of two pre-specified validation criteria of confirming similarity to the ONTARGET trial (HR 1.01 (95% CI: 0.94 to 1.09)) but not the second, because the CI did not include 1. Due to the much larger size of the observational study we analysed a randomly-selected subset of the cohort consistent with the trial size (n=17,687) which gave results consistent with ONTARGET: HR 0.97 (95% CI: 0.89 to 1.06): failure to meet the second pre-specified criteria is therefore likely due to an increase in power and thus narrower CIs, but subsequent results should be interpreted with this caveat. Results for secondary outcomes showed that ARB were associated with a lower risk than ACEi but 95% CIs overlapped with estimates from ONTARGET. Compared with ACEi, ARB were associated with an increased risk of loss of GFR or ESKD and doubling of creatinine.

### Extending analysis to the underrepresented ethnic groups

#### Primary composite outcome

Among Black, South Asian, and White patients, the primary composite outcome occurred in 2,568 (14.6%), 5,210 (16.9%) and 100,173 (19.1%) people, respectively. Event rates for Black patients were 20.5% and 20.2% per 5.5 person-years, 22.9% and 23.7% per 5.5 person-years for South Asian patients and for White patients, 25.3% and 26.4% per 5.5 person-years for ARB and ACEi users, respectively. For the comparison of ARB vs ACEi for the primary composite outcome there was no evidence of heterogeneity by ethnicity in the Cox model (*P_int_*=0.422). HR was 1.02 (95% CI: 0.93 to 1.12) for Black patients, HR 0.97 (95% CI: 0.91 to 1.03) for South Asian patients and HR 0.96 (95% CI: 0.95 to 0.98) for White patients. There was also no evidence of heterogeneity by ethnicity using the Poisson model to examine differences in incidence rates (*P_int_*=0.287) (Figure 2, Tables 2-3).

**Figure 2.**
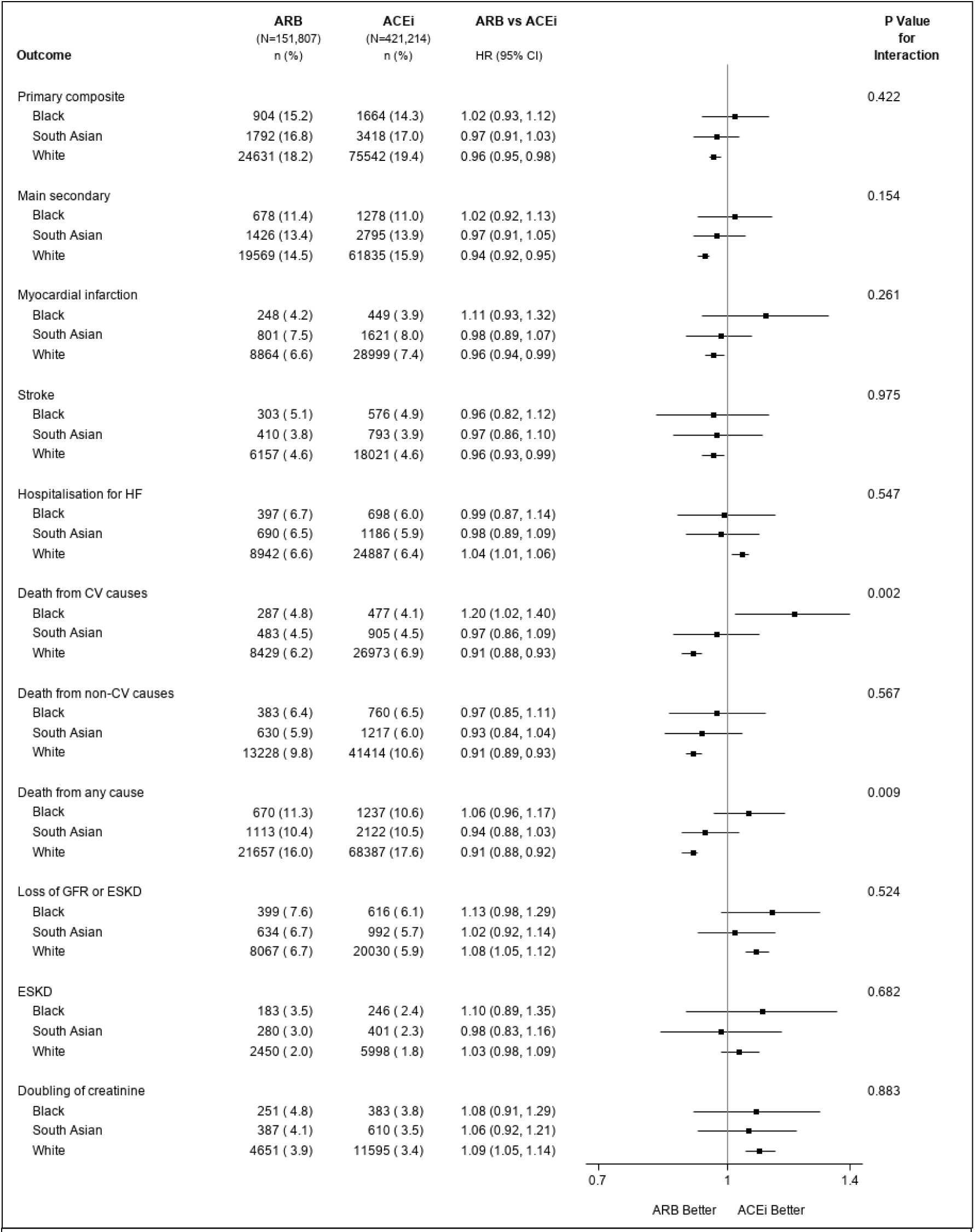
Treatment effect heterogeneity for the primary and secondary outcomes by ethnicity for extending analysis to underrepresented groups for ARB vs ACEi using a propensity-score—weighted analysis of trial-eligible patients in CPRD Aurum. Notes: N (%) = number of events (percent); ESKD: end-stage kidney disease; GFR: glomerular filtration rate. Primary composite outcome: death from cardiovascular causes, myocardial infarction, stroke, or hospitalisation for heart failure. Main secondary outcome: death from cardiovascular causes, myocardial infarction, or stroke. Loss of GFR or ESKD defined as: 50% reduction in estimated glomerular filtration ratio (eGFR), start of kidney replacement therapy (KRT) or eGFR<15ml/min/1.73m^2^. ESKD defined as: start of KRT or eGFR<15ml/min/1.73m^2^. P-value is test of interaction between ethnicity and treatment.

**Table 2.**
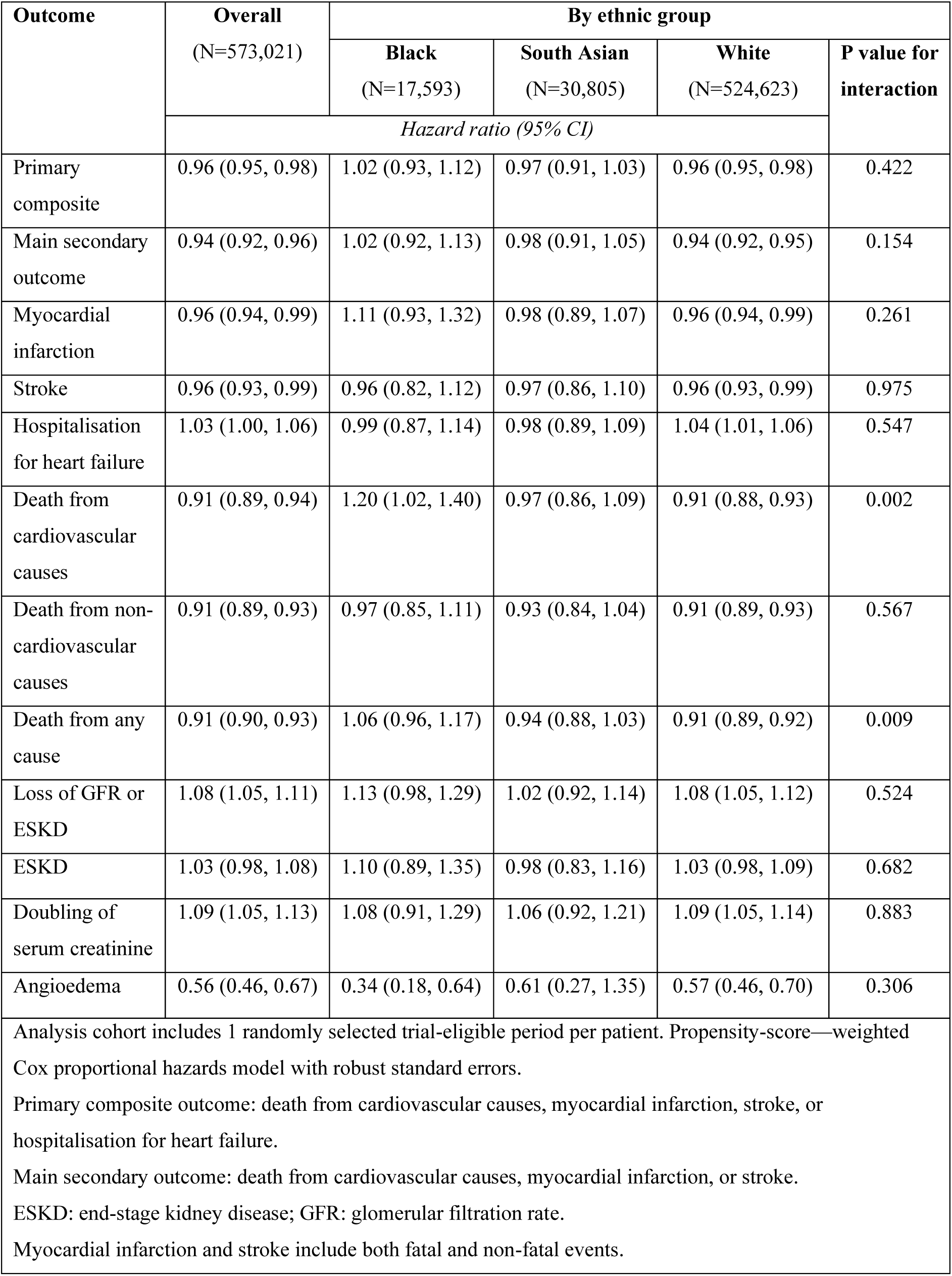
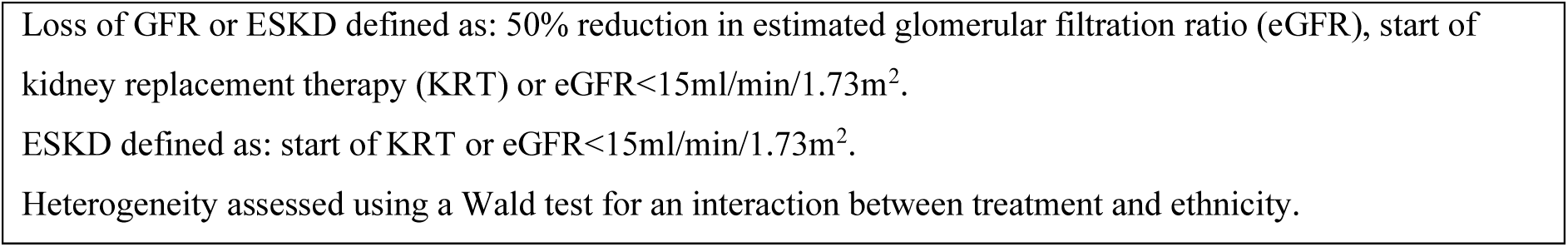
Results from extending analysis to underrepresented groups for ARB vs ACEi using a propensity-score—weighted analysis of trial-eligible patients in CPRD Aurum using a test for multiplicative heterogeneity.

**Table 3.**
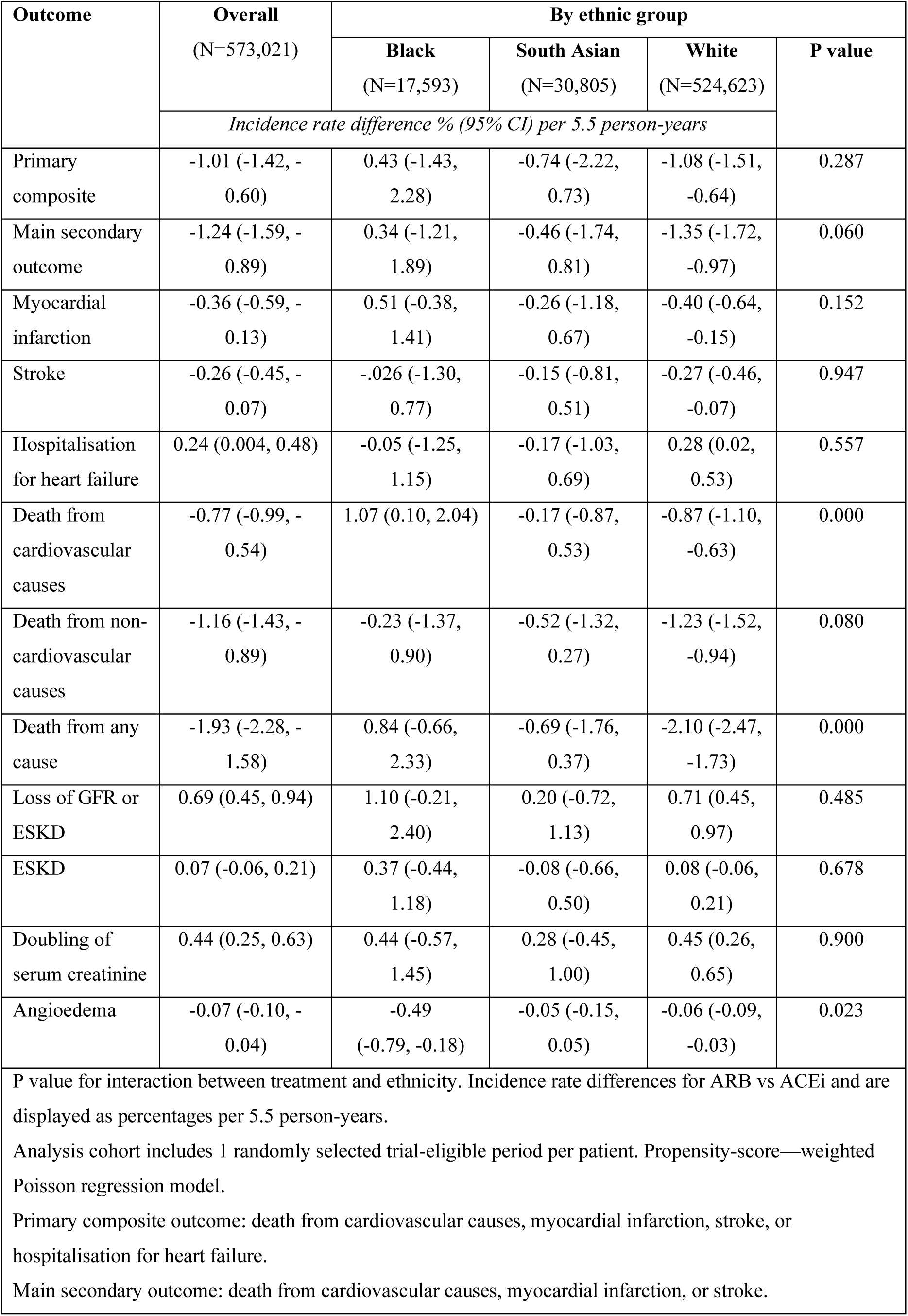
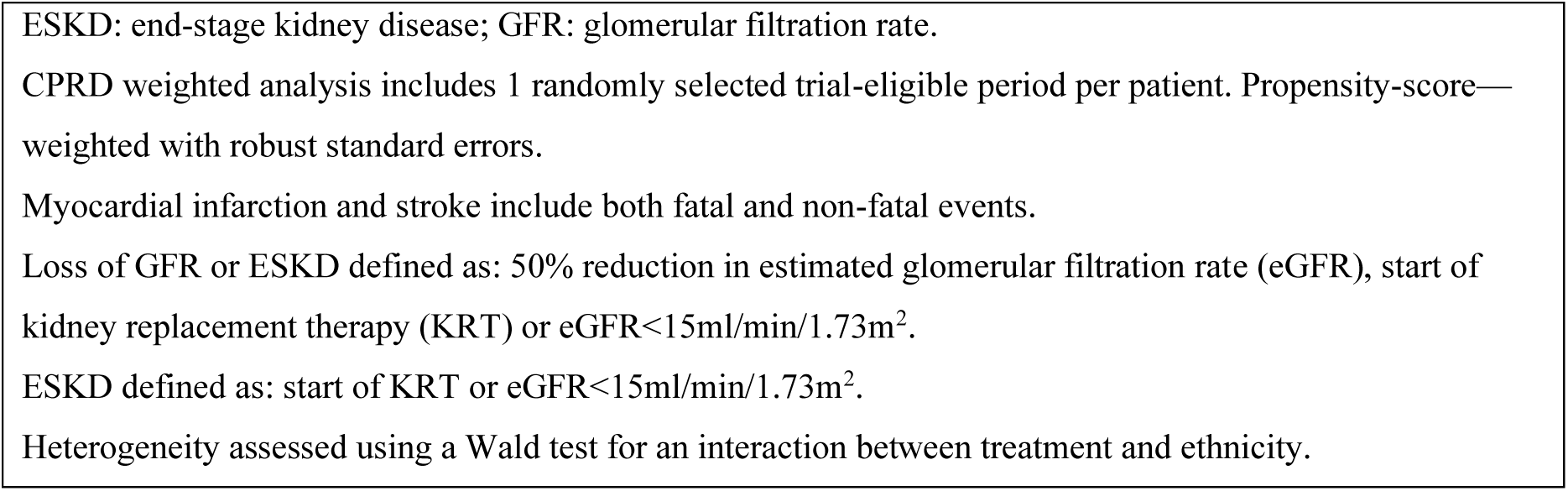
Results from extending analysis to underrepresented groups for ARB vs ACEi using a propensity-score—weighted analysis of trial-eligible patients in CPRD Aurum using a test for additive heterogeneity.

| Outcome | Overall<br>(N=573,021) | By ethnic group |  |  |  |
| --- | --- | --- | --- | --- | --- |
|  |  | Black<br>(N=17,593) | South Asian<br>(N=30,805) | White<br>(N=524,623) | P value |
|  | *Incidence rate difference % (95% CI) per 5.5 person-years* |  |  |  |  |
| Primary composite | -1.01 (-1.42, -0.60) | 0.43 (-1.43, 2.28) | -0.74 (-2.22, 0.73) | -1.08 (-1.51, -0.64) | 0.287 |
| Main secondary outcome | -1.24 (-1.59, -0.89) | 0.34 (-1.21, 1.89) | -0.46 (-1.74, 0.81) | -1.35 (-1.72, -0.97) | 0.060 |
| Myocardial infarction | -0.36 (-0.59, -0.13) | 0.51 (-0.38, 1.41) | -0.26 (-1.18, 0.67) | -0.40 (-0.64, -0.15) | 0.152 |
| Stroke | -0.26 (-0.45, -0.07) | -0.026 (-1.30, 0.77) | -0.15 (-0.81, 0.51) | -0.27 (-0.46, -0.07) | 0.947 |
| Hospitalisation for heart failure | 0.24 (0.004, 0.48) | -0.05 (-1.25, 1.15) | -0.17 (-1.03, 0.69) | 0.28 (0.02, 0.53) | 0.557 |
| Death from cardiovascular causes | -0.77 (-0.99, -0.54) | 1.07 (0.10, 2.04) | -0.17 (-0.87, 0.53) | -0.87 (-1.10, -0.63) | 0.000 |
| Death from non-cardiovascular causes | -1.16 (-1.43, -0.89) | -0.23 (-1.37, 0.90) | -0.52 (-1.32, 0.27) | -1.23 (-1.52, -0.94) | 0.080 |
| Death from any cause | -1.93 (-2.28, -1.58) | 0.84 (-0.66, 2.33) | -0.69 (-1.76, 0.37) | -2.10 (-2.47, -1.73) | 0.000 |
| Loss of GFR or ESKD | 0.69 (0.45, 0.94) | 1.10 (-0.21, 2.40) | 0.20 (-0.72, 1.13) | 0.71 (0.45, 0.97) | 0.485 |
| ESKD | 0.07 (-0.06, 0.21) | 0.37 (-0.44, 1.18) | -0.08 (-0.66, 0.50) | 0.08 (-0.06, 0.21) | 0.678 |
| Doubling of serum creatinine | 0.44 (0.25, 0.63) | 0.44 (-0.57, 1.45) | 0.28 (-0.45, 1.00) | 0.45 (0.26, 0.65) | 0.900 |
| Angioedema | -0.07 (-0.10, -0.04) | -0.49 (-0.79, -0.18) | -0.05 (-0.15, 0.05) | -0.06 (-0.09, -0.03) | 0.023 |
| P value for interaction between treatment and ethnicity. Incidence rate differences for ARB vs ACEi and are displayed as percentages per 5.5 person-years.<br>Analysis cohort includes 1 randomly selected trial-eligible period per patient. Propensity-score—weighted Poisson regression model.<br>Primary composite outcome: death from cardiovascular causes, myocardial infarction, stroke, or hospitalisation for heart failure.<br>Main secondary outcome: death from cardiovascular causes, myocardial infarction, or stroke. |  |  |  |  |  |

ESKD: end-stage kidney disease; GFR: glomerular filtration rate.
CPRD weighted analysis includes 1 randomly selected trial-eligible period per patient. Propensity-score—weighted with robust standard errors.
Myocardial infarction and stroke include both fatal and non-fatal events.
Loss of GFR or ESKD defined as: 50% reduction in estimated glomerular filtration rate (eGFR), start of kidney replacement therapy (KRT) or $\text{eGFR} < 15 \text{ ml/min/1.73m}^2$ .
ESKD defined as: start of KRT or $\text{eGFR} < 15 \text{ ml/min/1.73m}^2$ .
Heterogeneity assessed using a Wald test for an interaction between treatment and ethnicity.

#### Secondary outcomes

There was no evidence of treatment heterogeneity by ethnicity for the majority of secondary outcomes (Figure 2, Tables 2-3). However, there was evidence of heterogeneity for cardiovascular-related death (*P_int_* =0.002 and *P_int_*<0.001) and all-cause mortality (*P_int_*=0.009 and *P_int_* <0.001) on the multiplicative scale using the Cox model and on the additive scale for the difference in incidence rates, respectively. ARB were associated with reduced cardiovascular-related death for White patients (HR 0.91 (95% CI: 0.88 to 0.93); NNT 115 (95% CI: 91 to 159) for ARB vs ACEi) but associated with increased cardiovascular-related death for Black patients (HR 1.20 (95% CI: 1.02 to 1.40); NNH 93 (95% CI: 49 to 1000) for ARB vs ACEi). However for South Asian patients there was no evidence of a difference between ARB and ACEi treatment for risk of cardiovascular-related death (HR 0.97 (95% CI: 0.86 to 1.09)).

ARB were associated with reduced all-cause mortality compared with ACEi for White patients (HR 0.91 (95% CI: 0.89 to 0.92)) but no difference observed for Black (HR 1.06 (95% CI: 0.96 to 1.17)) or South Asian patients (HR 0.94 (95% CI: 0.88, 1.03) (Figure 2, Table 2). For the kidney outcomes, there was no significant heterogeneity by ethnicity (*P_int_*=0.485 to 0.900).

#### Angioedema

The overall incidence of angioedema recorded in our study population was 814 (0.14%) patients during the follow-up period of 5.5 years, with HR 0.56 (95% CI: 0.46 to 0.67) for ARB vs ACEi. Of these events, 35% occurred within the first 12 months. Over the total duration of follow-up (maximum 5.5 years), there was no evidence of heterogeneity on the multiplicative scale by ethnicity (*P_int_*=0.306) (Figure 3, Table 2). For ARB and ACEi, respectively, the angioedema rate was 0.33% and 0.82% in Black patients, 0.09% and 0.14% in South Asian patients and 0.13% and 0.19% in White patients per 5.5 person-years, with differences displayed in Table 3. Because of these differences in incidence rates, there was evidence of heterogeneity by ethnicity on the additive scale (*P_int_*=0.023). For ARB vs ACEi, NNT 204 (95% CI: 127 to 556) in Black patients and NNT 1667 (95% CI: 1111 to 3333) in White patients. There was no difference observed for South Asian patients.

**Figure 3.**
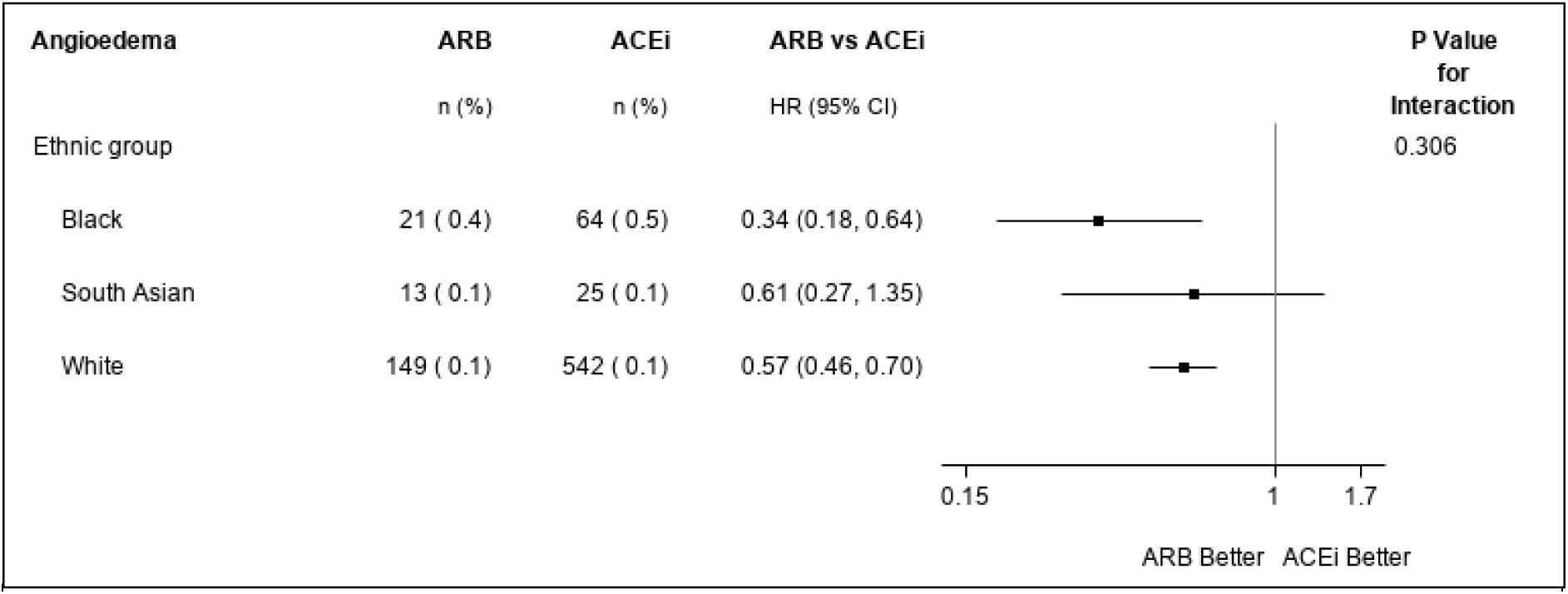
Treatment effect heterogeneity for the risk of angioedema by ethnicity for extending analysis to underrepresented groups for ARB vs ACEi using a propensity-score—weighted analysis of trial-eligible patients in CPRD Aurum. Notes: N (%)= number of events (percent). Total follow-up period is a maximum follow-up of 5.5 years with patients censored at death, transferred out of practice date or last collection date as in main analysis. Outcome assessed using propensity-score—weighted Cox proportional hazards model. P-value is test of interaction between ethnicity and treatment.

#### Blood pressure

South Asian patients had the lowest blood pressure at baseline, with Black patients having the highest (supplementary Table S4-S6). On average, Black patients had approximately 16 recorded blood pressure measurements within a maximum of 5.5 years follow-up, compared with 14 for White patients and 15 for South Asian patients. South Asian and White individuals starting an ACEi experienced a greater fall in systolic blood pressure than those starting ARB; there was no difference in blood pressure between ARB and ACEi for Black patients (Figure 4). The biggest decrease in BP was observed in the first six weeks of the trial-eligible period. Over 4 years of follow-up, mean systolic blood pressure was reduced by 5, 5.5 and 8 mm Hg among Black, South Asian, and White patients, respectively.

**Figure 4.**
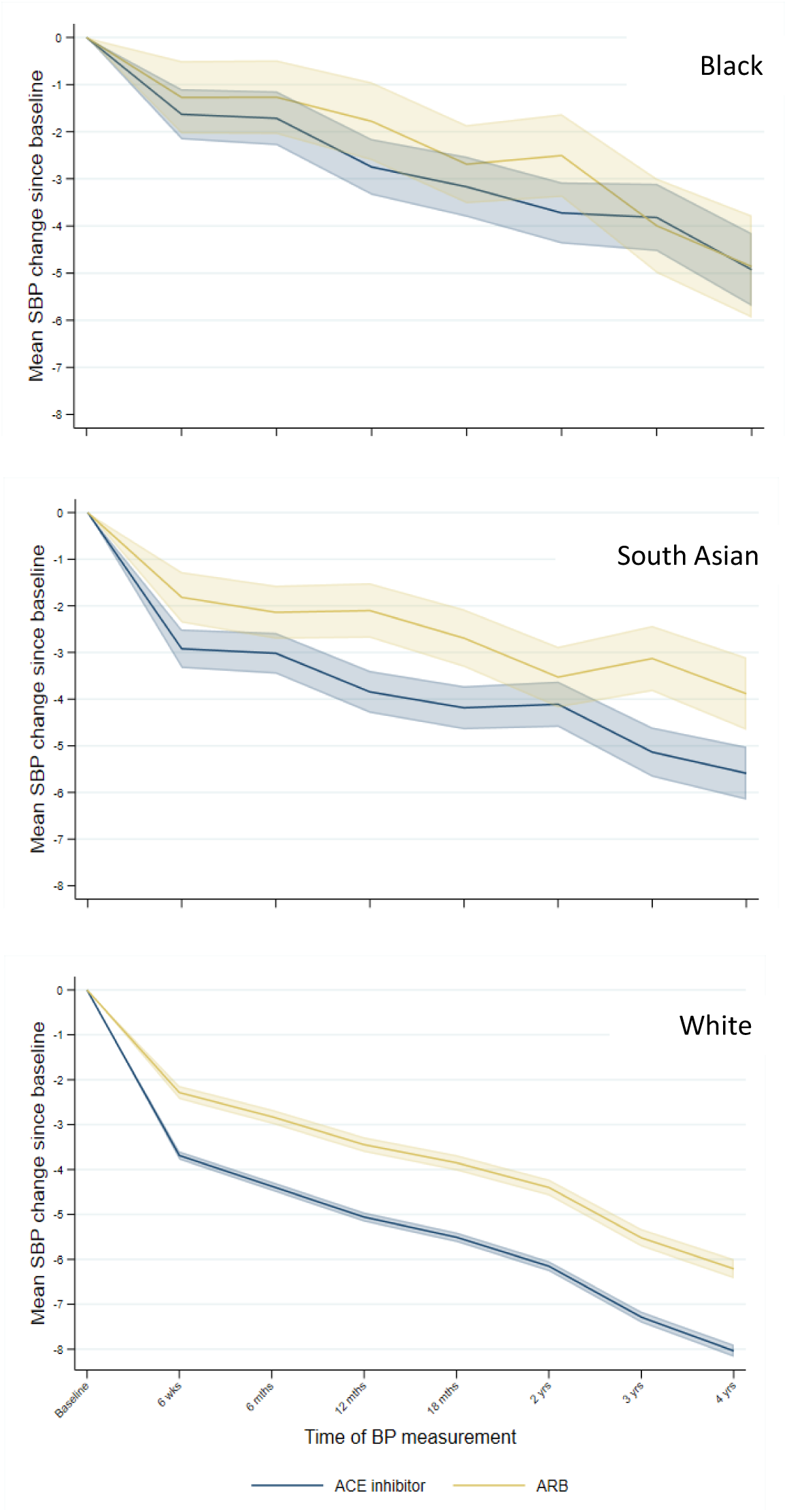
Changes in systolic blood pressure by treatment and ethnic group (mm Hg) with 95% confidence intervals. Notes: BP: blood pressure; SBP: systolic blood pressure. Baseline is closest measurement taken prior to start of trial-eligible period within 2 years.

### Sensitivity analyses

Results were consistent with the main analysis 1) when we restricted to follow-up time on the original treatment (on-treatment analysis) (supplementary Table S8); 2) when we used multiple imputation of missing covariates for variables included in the propensity score model to assess the bias introduced from a complete case analysis (supplementary Table S9). When extending analysis to underrepresented ethnic groups, assessing the impact of the 2011 treatment recommendation by restricting the cohort to trial-eligible periods prior to 2011 also gave consistent results (supplementary Figure S3).

## Discussion

### Principal findings

In this observational study with reference trial emulation reflecting current routine care in England, we successfully emulated a landmark randomised trial before extending analysis to South Asian and Black patients. We observed that patients treated with ARB and ACEi had similar risks of most cardiovascular outcomes, with no evidence of heterogeneity by ethnicity when analysed as hazard ratios or absolute incidence rate differences. For the outcome of cardiovascular death, ARB was associated with overall reduced risk compared to ACEi, but findings differed by ethnicity. Over the 5.5 year study period, Black people had 1.07% more cardiovascular deaths when treated with ARB compared with ACEi (NNH 93 (95% CI: 49 to 1000) for ARB vs ACEi). White people had fewer cardiovascular-related death outcomes when treated with ARB compared with ACEi (NNT 115 (95% CI: 91 to 159) for ARB vs ACEi). Outcomes were the same with both drugs for South Asian individuals. For the outcome of death from all causes, ARB were associated with decreased risk for White patients, with no difference in treatment effectiveness for Black and South Asian patients. For kidney outcomes, overall, treatment with ARB was associated with increased risk of loss of GFR or ESKD and doubling of creatinine, with no evidence of heterogeneity by ethnicity. ARB were associated with 44% less angioedema than ACEi overall, with no evidence of heterogeneity using a Cox regression model. However, incidence rates of angioedema were more common in Black patients than in White or South Asian patients, leading to heterogeneity on the additive scale: for ARB vs ACEi, the NNT for Black patients, over the 5.5 year study period, was 204 (95% CI: 127 to 556) whereas for White patients it was 1667 (95% CI: 1111 to 3333), with no difference observed for South Asian patients.

Black patients had highest blood pressure measurements at baseline. However, we observed greater levels of blood pressure reduction after treatment initiation for White individuals compared to Black and South Asian ethnic groups.

### Strengths and limitations

Our large sample size and use of a reference trial emulation approach increases confidence in our findings of comparative effectiveness of ARB and ACEi in preventing cardiovascular outcomes in Black and South Asian populations, who are often underrepresented in trials. Unlike traditional RCTs, this study design enables us to explore effects in large, diverse samples with the possibility to identify subgroup effects which may have been missed in trials due to the limited sample size. To our knowledge this is also the first study exploring the risk of angioedema associated with ARB and ACEi treatment use among a large ethnically diverse population in the UK. However our analysis does have limitations. Firstly, our analysis did not meet both pre-specified criteria of trial replicability in our benchmarking analysis but this is likely due to an increase in power and narrower confidence intervals. Secondly by repeating all the outcomes of the ONTARGET trial there is a risk of chance findings due to multiple testing. There could be residual confounding from the clinicians decision to prescribe ACEi or ARB in response to uncaptured information about the type or severity of condition being treated. It is likely that this bias would lead to increased risk observed among patients prescribed ACEi since this is the recommended first line treatment for patients with heart failure with reduced ejection fraction.[39] Indeed, this may underly the apparent reduced risk of death from cardiovascular and all causes seen in the study overall and among White participants. However the results for cardiovascular mortality among Black participants are in contrast to this presumed source of bias. The exclusion of patients with missing ethnicity could introduce bias into our results; however, ethnicity is well-captured using combined CPRD Aurum and HES data, so only 1.6% of patients were excluded. After imputing missing values for baseline blood pressure and creatinine, the association between ARB use and cardiovascular-death in Black patients was reduced in magnitude on the relative scale but results remained consistent with an increased risk of cardiovascular-death for Black patients and a reduced risk of cardiovascular-death for White patients. In agreement with other studies assessing the incidence of angioedema associated with ACEi use, incidence was low.[19, 20, 40] However, as we assessed the risk of angioedema using only events reported in primary care we may have underestimated the true event rate. Low power for this outcome means these results must be interpreted with caution. In addition recording of angioedema in Black patients, particularly those taking ACEi, could be influenced by increased recognition and thus differential misclassification by clinicians aware of an association.

### Comparison with literature

Our results support the generalisability of the ONTARGET trial results[23] to ethnic minority populations, but raise the question of whether, for the prevention of cardiovascular death, treatment with ACEi might be associated with fewer events in people who are Black and ARB in people who are white. Unlike most areas of clinical medicine, variations in drug effect by ethnicity are biologically plausible: differences in the renin-angiotensin system by ethnicity underly the current UK hypertension treatment recommendations[14] and an increase in bradykinin which occurs with ACEi treatment has been proposed as contributing to the therapeutic effects of these drugs.[17] Randomised evidence of different treatment effects among people of different ethnicities is limited. A 2014 Cochrane meta-analysis of head-to-head trials in people with hypertension found no differences between ARB and ACEi, but did not include a subgroup analysis by ethnicity.[41] A large observational study across multiple databases, comparing ARB with ACEi, found no differences for any cardiovascular outcome or the cardiovascular composite; no information on ethnicity was reported.[23, 42]

In terms of angioedema, despite a subgroup analysis of ALLHAT indicating increased incidence among Black users of ACEi use compared to those who were non-Black, the trial did not include a direct comparison between ARB and ACEi.[20] Black individuals had an a 3- to 4-fold increase in the risk of angioedema compared with non-Black individuals, regardless of treatment. We also observed a 2- to 3-fold increase in risk for ACEi compared with ARB which is comparable to the relative risk of 3.6 for ACEi compared with other anti-hypertensives in a study of US Veterans.[43] Our work is the first to quantify the interaction with ethnicity and to report NNH by ethnicity, and the first study of a population outside the US.[43, 44]

### Interpretations and conclusions

In conclusion, we observed similar treatment effects of ARB and ACEi in preventing cardiovascular outcomes in high-risk South Asian and Black individuals using routinely-collected data from the UK, consistent with the ONTARGET trial findings.

Without replication, it is uncertain to what extent our finding of differences in outcomes for ARB vs ACEi by ethnicity should influence guidelines. Differences in outcomes for ARB vs ACEi for the primary composite outcome were not associated with a statistically significant interaction term (*P_int_*=0.422). However, for the outcome death from cardiovascular causes, and the highly credible outcome of all-cause mortality, there was a highly statistically significant interaction (*P_int_*=0.002 and *P_int_*<0.001) favouring ACEi for Black individuals, ARB for White individuals, and with no difference for South Asian individuals. In Black individuals, we observed 93 patients were required to be treated with an ARB instead of an ACEi for one additional patient to experience cardiovascular death. This NNH, is smaller by a factor of 2 than the NNT for treatment with an ARB instead of an ACEi to prevent one additional angioedema event (204). Therefore, even if events were of equal severity, alteration of current UK recommendations to choose ARB over ACEi for Black individuals should be considered. Whether long-term outcomes of ACEi or ARB have different effects in people of different ethnicities could be addressed in a large nationwide pragmatic trial with randomisation at treatment initiation in primary care.

## Data Availability

No additional data available

## Contributors

All authors contributed to the conceptualisation, study question and design. PB, LT, KW, EW, AW, AS, CL, CC, JM and EP contributed to the statistical analysis decisions of this study. PB completed formal analysis of this study and wrote the original draft of this manuscript under the supervision of LT, KW and MC. PB, LT, KW, EW, AW, AS, CL, EP had access to the data and data reported in this manuscript was verified by LT and KW. All authors contributed to the writing, review and editing of the submitted manuscript.

## Ethical approval

Ethical approval has been granted by the London School of Hygiene & Tropical Medicine Ethics Committee (Ref: 22658). The study has been approved by the Independent Scientific Advisory Committee of the UK Medicines and Healthcare Products Regulatory Agency (protocol no. 20_012). Access to the individual patient data from the ONTARGET trial was obtained by the trial investigators and complies with institutional review board approved informed consent forms provided by the individuals from whom the data were collected.

## Data sharing

No additional data available

## Transparency declaration

The manuscript’s guarantor (PB) affirms that this manuscript is an honest, accurate, and transparent account of the study being reported; that no important aspects of the study have been omitted; and that any discrepancies from the study as planned have been explained.

## Dissemination

We are not able to disseminate the results of this research directly to study participants as the data used were anonymised.

