## Supplementary figures and images for "Comparative effectiveness of ARB and ACEi for cardiovascular outcomes and risk of angioedema among different ethnic groups in England: an analysis in the UK Clinical Practice Research Datalink with emulation of a reference trial (ONTARGET)"

### Supplementary Figure S1

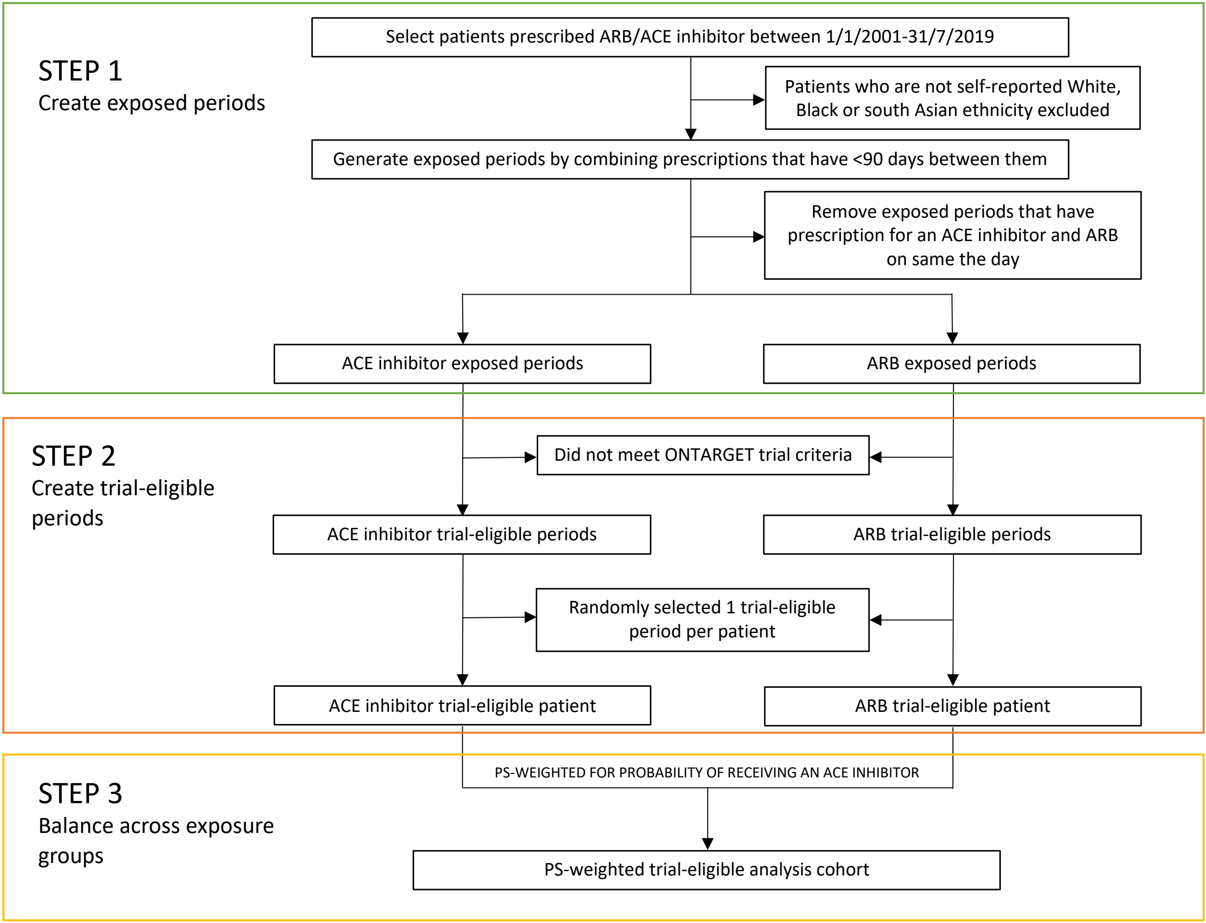


**Supplementary Figure S1.** Steps to define analysis cohort.

PS=propensity score
