## Supplementary Figure S2 for "Comparative effectiveness of ARB and ACEi for cardiovascular outcomes and risk of angioedema among different ethnic groups in England: an analysis in the UK Clinical Practice Research Datalink with emulation of a reference trial (ONTARGET)"

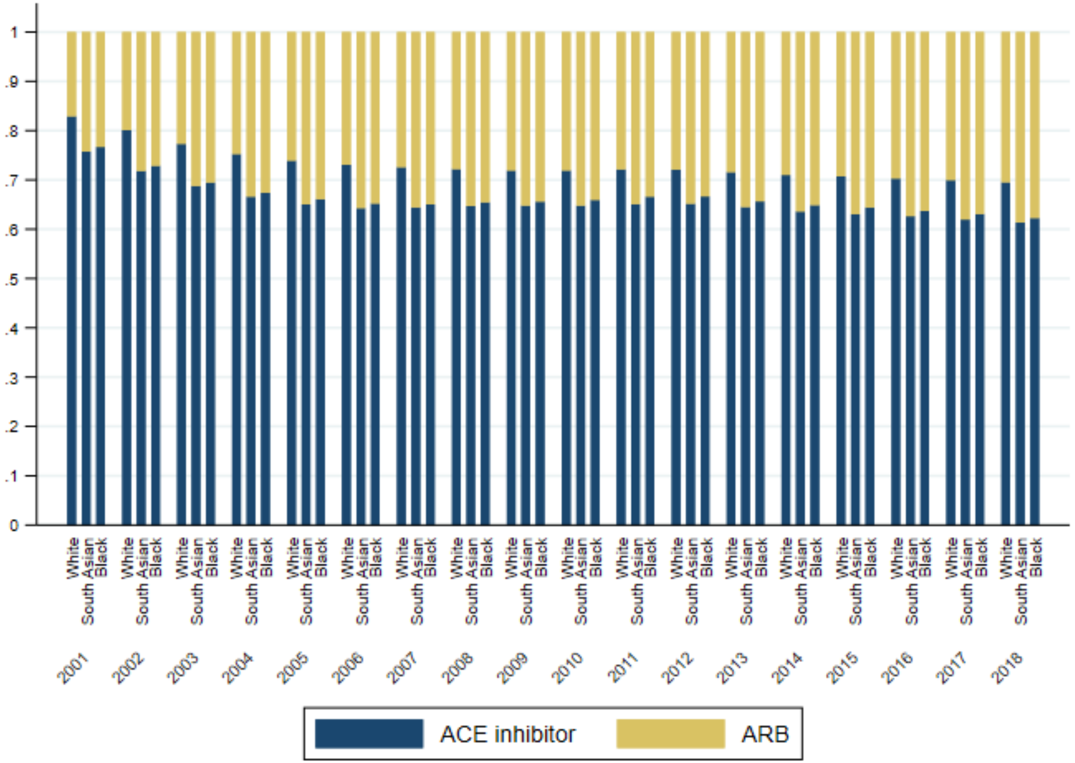


**Supplementary Figure S2.** Proportion of ARB and ACE inhibitor prescriptions prescribed each year out of total number prescribed within each ethnic group.
