## Supplementary Figure S3 for "Comparative effectiveness of ARB and ACEi for cardiovascular outcomes and risk of angioedema among different ethnic groups in England: an analysis in the UK Clinical Practice Research Datalink with emulation of a reference trial (ONTARGET)"

| Primary outcome (sensitivity) | ARB<br>n (%) | ACEi<br>n (%) | ARB vs ACEi<br>HR (95% CI) | P Value for Interaction |
|---|---|---|---|---|
| On-treatment approach | | | | 0.581 |
| Black | 843 (14.2) | 1607 (13.8) | 1.00 (0.91, 1.10) | |
| South Asian | 1682 (15.8) | 3323 (16.5) | 0.95 (0.89, 1.02) | |
| White | 23675 (17.5) | 74707 (19.2) | 0.95 (0.94, 0.97) | |
| Eligible period <2011 | | | | 0.232 |
| Black | 425 (16.4) | 850 (14.9) | 1.04 (0.92, 1.19) | |
| South Asian | 803 (18.3) | 1649 (18.0) | 0.98 (0.90, 1.08) | |
| White | 13325 (19.1) | 43996 (20.5) | 0.94 (0.92, 0.96) | |

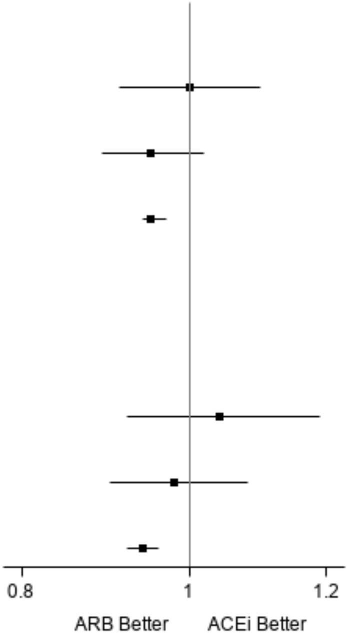


**Supplementary Figure S3.** Forest plot of sensitivity analysis for extending analysis to underrepresented groups using a propensity-score—weighted analysis for ARB vs ACEi use for primary composite outcome. Primary composite outcome is cardiovascular related death, myocardial infarction, stroke, or hospitalisation for heart failure. On-treatment approach censored at treatment discontinuation (i.e., treatment gap of >90 days), switch treatment or start of dual use +60 days. Multiple imputation of missing baseline blood pressure and creatinine using chained equations. Eligible period <2011 is analysis restricted to start of trial-eligible periods prior to 2011. P value is test for heterogeneity using an term interaction between treatment and ethnicity in the Cox proportional hazards model.
