## Supplementary Table S1 for "Comparative effectiveness of ARB and ACEi for cardiovascular outcomes and risk of angioedema among different ethnic groups in England: an analysis in the UK Clinical Practice Research Datalink with emulation of a reference trial (ONTARGET)"

| **Supplementary Table S1.** Key design aspects of the reference trial (ONTARGET) and the reference trial emulation using CPRD Aurum data | | |
| --- | --- | --- |
| **Protocol component** | **Reference trial (ONTARGET)** | **Emulation in CPRD Aurum** |
| Eligibility criteria | Patients aged ≥55 years with coronary artery, peripheral artery or cerebrovascular disease or high-risk diabetes with end organ damage recruited up to 2004. No restriction on previous ACEi/ARB use except must be able to discontinue use. | Black, South Asian or White patients with a prescription for an ACEi or ARB between 2001-01-2001 to 2019-07-31, eligible for HES linkage, aged ≥55 years with coronary artery, peripheral vascular, or cerebrovascular disease or high-risk diabetes. |
| Treatment strategies | Patients entered 3-week single blind run-in period to check compliance then randomised to one of three trial arms: ramipril 10 mg + telmisartan placebo, telmisartan 80 mg + ramipril placebo or ramipril 10 mg + telmisartan 80 mg. | Exposure groups defined by prescriptions for ARB or ACEi. Exposed periods will be continuous courses of therapy. A new exposed period will begin when a prescription gap of >90 days occurs. |
| Assignment procedures | Randomly assigned and received placebo for other drug so unaware which arm assigned to | Based on prescriptions received. Patient can contribute to both exposure groups at different timepoints |
| Follow-up period | Follow-up started at randomisation and ended at primary event, death, loss to follow-up or end of study. Close out was planned in July 2007. | Follow-up starts at start of trial-eligible period where exposure period meets trial inclusion/exclusion criteria. Ends at the earliest of: outcome of interest, death, transferred out of practice date, or last data collection from the general practice. If these dates do not occur the patient will be censored after 5.5 years of follow-up |
| Outcome | Primary composite of: cardiovascular death, MI, stroke, hospitalisation for heart failure | As in ONTARGET, defined using ICD10, Read codes and death registries from ONS. |
| Analysis plan | Primary analysis under time-to-event counting first occurrence of any component of the composite outcome using Cox proportional hazards model. Intention-to-treat as main analysis | Analysis conducted on one randomly selected trial eligible period per patient. Balance of covariates obtained by propensity score weighting for probability of receiving an ACEi. Weighting as opposed to matching to increase sample size and diversity of cohort to enable inferences to be extended to underrepresented groups of Black and South Asian individuals. Cox proportional hazards model used for primary analysis. |
| *Restricted to those of Black, South Asian and White ethnicity as other and mixed ethnic group had insufficient numbers. | | |
