## Supplementary Table S2 for "Comparative effectiveness of ARB and ACEi for cardiovascular outcomes and risk of angioedema among different ethnic groups in England: an analysis in the UK Clinical Practice Research Datalink with emulation of a reference trial (ONTARGET)"

| **Supplementary Table S2** List of variables considered and included in propensity-score model for balancing characteristics between exposure groups | | | |
| --- | --- | --- | --- |
| **Potential confounders** | **Selected into propensity-score model** | **Missing data** | **Reason for omitting from PS model** |
| Stroke/TIA | ✓ | - |  |
| Peripheral artery disease | ✓ | - |  |
| Coronary artery disease | ✓ | - |  |
| Diabetes | ✓ | - |  |
| High-risk diabetes | ✓ | - |  |
| Age (years) | ✓ | - |  |
| Sex | ✓ | - |  |
| Ethnicity | ✓ | - |  |
| BMI | ✓ | 7% |  |
| SBP | ✓ | 22% |  |
| DBP | ✓ | 22% |  |
| Creatinine | ✓ | 19% |  |
| Index of Multiple Deprivation (IMD) | ✓ | 0.1% |  |
| Smoke status | ✓ | 2.4% |  |
| Alcohol use |  | 16% | Missing data |
| Statin use | ✓ | - |  |
| Nitrate use | ✓ | - |  |
| Diabetic treatment use | ✓ | - |  |
| Diuretic use | ✓ | - |  |
| CCB use | ✓ | - |  |
| Betablocker use | ✓ | - |  |
| Aspirin use | ✓ | - |  |
| Antiplatelet use | ✓ | - |  |
| Digoxin use |  | - | Insufficient number of events |
| Anticoagulant use | ✓ | - |  |
| Alpha-blocker use | ✓ | - |  |
| No. of hospital admissions within 6 months prior | ✓ | - |  |
| No. of GP appointments within 6 months prior | ✓ | - |  |
| Year of start of eligible period | ✓ | - |  |
| Time since first eligible period (days) | ✓ | - |  |
| No. of previous ACE inhibitor eligible periods | ✓ | - |  |
| No. of previous ARB eligible periods | ✓ | - |  |
| Notes: TIA: transient ischaemic attack; BMI: body-mass index; SBP: systolic blood pressure; DBP: diastolic blood pressure.  Variables are measured at start of trial-eligible period or before.  Peripheral artery disease includes limb bypass surgery or angioplasty, limb/foot amputation, or intermittent claudication.  Coronary artery disease includes previous MI, angina, coronary angioplasty, or CABG.  SBP and DBP are measured within 6 months prior to start of trial-eligible period.  Medication use is within 3 months prior to start of trial-eligible period.  SBP and DBP had 22% missing data but this variable was included as believed to be an important confounder and can be assumed to be MAR. Creatinine was included as a binary indicator for missing and non-missing and an additional variable where missing values were imputed as the mean. Alcohol was omitted due to missing data >10% and reason to not assume to be MAR. If balance was unachieved for this variable after propensity-score—weighting it would be considered in the model with a missing value category.  Balance after weighting was assessed for all variables listed including those not included in the propensity-score model. | | | |
