## Supplementary Table S3 for "Comparative effectiveness of ARB and ACEi for cardiovascular outcomes and risk of angioedema among different ethnic groups in England: an analysis in the UK Clinical Practice Research Datalink with emulation of a reference trial (ONTARGET)"

| **Supplementary Table S3** Baseline characteristics and standardised differences of trial-eligible patients after applying trial criteria included in propensity-score—weighted analysis before and after weighting for the reference trial emulation | | | | | |
| --- | --- | --- | --- | --- | --- |
| **Characteristic** | **Before weighting** | | **After weighting** | | |
|  | **ARB**  N=151,807 | **ACEi**  N=421,214 | **ARB**  N=554,609 | **ACEi**  N=562,686 | **SMD** |
| **Age (year) – mean (SD)** | 71.3 (9.2) | 70.9 (9.4) | 554609 (71.1) | 562686 (71.0) | 0.007 |
| **Systolic BP (mmHg) – mean (SD)** | 144.0 (20.2) | 143.6 (20.2) | 144.7 (20.6) | 144.1 (20.3) | 0.031 |
| **Diastolic BP (mmHg) – mean (SD)** | 78.8 (10.9) | 79.1 (11.0) | 79.3 (11.0) | 79.2 (11.0) | 0.015 |
| **Body-mass index – mean (SD)** | 29.2 (5.8) | 28.8 (5.8) | 28.9 (5.7) | 28.9 (5.8) | 0.010 |
| **Creatinine (μmol/l) – mean (SD)** | 94.1 (29.5) | 92.9 (27.3) | 93.8 (28.6) | 93.5 (28.1) | 0.012 |
| **Female sex – no. (%)** | 82849 (54.6) | 202511 (48.1) | 278912 (50.3) | 280169 (49.8) | 0.010 |
| **Ethnic group – no. (%)** |  |  |  |  |  |
| Black | 5947 (3.9) | 11646 (2.8) | 17144 (3.1) | 17371 (3.1) | 0.000 |
| South Asian | 10668 (7.0) | 20137 (4.8) | 30176 (5.4) | 30309 (5.4) | 0.002 |
| White | 135192 (89.1) | 389431 (92.5) | 507289 (91.5) | 515006 (91.5) | 0.002 |
| **Clinical history – no. (%)** |  |  |  |  |  |
| CAD^a^ | 105969 (69.8) | 298697 (70.9) | 389536 (70.2) | 396723 (70.5) | 0.006 |
| Cerebrovascular disease^b^ | 16071 (10.6) | 44821 (10.6) | 58605 (10.6) | 59511 (10.6) | 0.000 |
| PAD^c^ | 14158 (9.3) | 38958 (9.3) | 51159 (9.2) | 52018 (9.2) | 0.001 |
| Diabetes | 93709 (61.7) | 250030 (59.4) | 333282 (60.1) | 337295 (59.9) | 0.003 |
| High-risk diabetes^d^ | 76297 (50.3) | 197471 (46.9) | 270032 (48.7) | 270292 (48.0) | 0.013 |
| **Smoking status – no. (%)** |  |  |  |  |  |
| Non-smoker | 44137 (29.1) | 113688 (27.0) | 153008 (27.6) | 155056 (27.6) | 0.000 |
| Current smoker | 35111 (23.1) | 111998 (26.6) | 141002 (25.4) | 144500 (25.7) | 0.007 |
| Past smoker | 72559 (47.8) | 195528 (46.4) | 260599 (47.0) | 263129 (46.8) | 0.006 |
| **Alcohol drinker – no. (%)** |  |  |  |  |  |
| Yes | 92697 (61.1) | 261221 (62.0) | 344124 (62.1) | 345638 (61.4) | 0.014 |
| No | 45266 (29.8) | 119770 (28.4) | 157051 (28.3) | 164181 (29.2) | 0.023 |
| Unknown | 13844 (9.1) | 40223 (9.6) | 53433 (9.6) | 52867 (9.4) |  |
| **Medication^e^ – no. (%)** |  |  |  |  |  |
| Alpha-blocker | 17089 (11.3) | 38166 (9.1) | 56547 (10.2) | 54990 (9.8) | 0.001 |
| Oral anticoagulant agent | 13055 (8.6) | 34986 (8.3) | 47457 (8.6) | 47408 (8.4) | 0.009 |
| Antiplatelet agent | 13365 (8.8) | 40980 (9.7) | 53907 (9.7) | 53865 (9.6) | 0.007 |
| Aspirin | 50393 (33.2) | 148837 (35.3) | 196084 (35.4) | 196731 (35.0) | 0.003 |
| Beta-blocker | 47734 (31.4) | 135829 (32.3) | 181804 (32.8) | 181431 (32.2) | 0.009 |
| Calcium-channel blocker | 52535 (34.6) | 135915 (32.3) | 189502 (34.2) | 186885 (33.2) | 0.010 |
| Digoxin | 5430 (3.6) | 16976 (4.0) | 20687 (3.7) | 22649 (4.0) | 0.015 |
| Diuretics | 63949 (42.1) | 163355 (38.8) | 228374 (41.2) | 225328 (40.1) | 0.015 |
| Diabetic treatment | 38321 (25.2) | 101647 (24.1) | 138112 (24.9) | 138250 (24.6) | 0.001 |
| Nitrates | 14102 (9.3) | 43919 (10.4) | 57489 (10.4) | 57559 (10.2) | 0.007 |
| Statins | 79774 (52.6) | 225469 (53.5) | 296624 (53.5) | 299874 (53.3) | 0.002 |
| **Time-related variables – mean (SD)** |  |  |  |  |  |
| Time since trial-eligible period | 441.8 (1001.9) | 269.9 (760.8) | 354.8 (906.2) | 312.7 (830.7) | 0.006 |
| Number of prior ARB eligible periods | 0.9 (1.3) | 0.1 (0.5) | 0.4 (0.9) | 0.3 (0.9) | 0.024 |
| Number of prior ACEi eligible periods | 1.4 (2.5) | 1.5 (2.4) | 1.5 (2.5) | 1.5 (2.4) | 0.029 |
| Calendar year | 2011 (5.4) | 2010 (5.3) | 2010 (5.4) | 2010 (5.3) | 0.008 |
| **Healthcare utilisation^f^ - mean (SD)** |  |  |  |  |  |
| Number of GP appointments | 5.8 (23.2) | 9.9 (28.5) | 8.5 (28.2) | 8.9 (27.1) | 0.001 |
| Number of hospital admissions | 2.9 (10.7) | 3.1 (10.4) | 3.3 (11.1) | 3.2 (10.8) | 0.024 |
| **Index of multiple deprivation – no. (%)** |  |  |  |  |  |
| 1 (least) | 30758 (20.3) | 81277 (19.3) | 107889 (19.5) | 109527 (19.5) | 0.002 |
| 2 | 32055 (21.1) | 86909 (20.6) | 114994 (20.7) | 116757 (20.8) | 0.001 |
| 3 | 30044 (19.8) | 82898 (19.7) | 109518 (19.8) | 110932 (19.7) | 0.002 |
| 4 | 30343 (20.0) | 85793 (20.4) | 112392 (20.3) | 114106 (20.3) | 0.000 |
| 5 (most) | 28607 (18.8) | 84337 (20.0) | 109817 (19.8) | 111365 (19.8) | 0.002 |
| N= number of patients; no. (%)=number (percent); SD= standard deviation; SMD=standardised mean difference; BP= blood pressure; CAD=coronary artery disease; PAD=peripheral artery disease  Post-weighting N displays weighted distribution of number of patients in the two exposure groups.  Propensity-score weights are unstabilized inverse probability weights.  One third of ONTARGET participants received both ramipril plus telmisartan.  ^a^ Includes diagnosis of: MI at least 2 days prior, angina at least 30 days prior, angioplasty at least 30 days prior, CABG at least 4 years prior  ^b^ Includes diagnosis of: stroke/TIA  ^c^ Includes diagnosis of: limb bypass surgery, limb/foot amputation, intermittent claudication  ^d^ Includes DM with: retinopathy, neuropathy, chronic kidney disease, proteinuria or other complication  ^e^ Within 3 months prior to eligible start date. Antiplatelet agent= clopidogrel/ticlopidine  ^f^ Within 6 months prior to eligible start date. | | | | | |
