## Supplementary Table S4 for "Comparative effectiveness of ARB and ACEi for cardiovascular outcomes and risk of angioedema among different ethnic groups in England: an analysis in the UK Clinical Practice Research Datalink with emulation of a reference trial (ONTARGET)"

| **Supplementary Table S4** Baseline characteristics and standardised differences of trial-eligible Black patients after applying trial criteria included in propensity-score—weighted analysis before and after weighting for extending analysis to underrepresented groups | | | | | |
| --- | --- | --- | --- | --- | --- |
| **Characteristic** | **Black ethnic group** | | | | |
|  | **Before weighting** | | **After weighting** | | |
|  | **ARB**  N=11,646 | **ACEi**  N=5,947 | **ARB**  N=17,144 | **ACEi**  N=17,371 | **SMD** |
| **Age (year) – mean (SD)** | 68.9 (8.9) | 68.5 (8.9) | 68.7 (8.9) | 68.7 (8.9) | 0.001 |
| **Systolic BP (mmHg) – mean (SD)** | 144.8 (19.3) | 144.3 (19.0) | 145.1 (19.6) | 144.8 (19.1) | 0.016 |
| **Diastolic BP (mmHg) – mean (SD)** | 80.2 (10.8) | 80.7 (10.8) | 80.7 (10.8) | 80.6 (10.8) | 0.003 |
| **Body-mass index – mean (SD)** | 30.5 (5.9) | 29.8 (5.7) | 30.2 (5.8) | 30.1 (5.7) | 0.016 |
| **Creatinine (μmol/l) – mean (SD)** | 99.0 (32.5) | 97.0 (29.2) | 98.2 (31.1) | 98.0 (30.6) | 0.005 |
| **Female sex – no. (%)** | 3439 (57.8) | 5982 (51.4) | 9247 (53.9) | 9314 (53.6) | 0.006 |
| **Clinical history – no. (%)** |  |  |  |  |  |
| CAD^a^ | 3883 (65.3) | 7812 (67.1) | 11318 (66.0) | 11511 (66.3) | 0.005 |
| Cerebrovascular disease^b^ | 586 (9.9) | 1169 (10.0) | 1736 (10.1) | 1738 (10.0) | 0.004 |
| PAD^c^ | 551 (9.3) | 1029 (8.8) | 1539 (9.0) | 1550 (8.9) | 0.002 |
| Diabetes | 4623 (77.7) | 9114 (78.3) | 13397 (78.2) | 13575 (78.1) | 0.000 |
| High-risk diabetes^d^ | 3537 (59.5) | 6763 (58.1) | 10159 (59.3) | 10249 (59.0) | 0.005 |
| **Smoking status – no. (%)** |  |  |  |  |  |
| Non-smoker | 2489 (41.9) | 4789 (41.1) | 7080 (41.3) | 7165 (41.2) | 0.001 |
| Current smoker | 1186 (19.9) | 2709 (23.3) | 3787 (22.1) | 3858 (22.2) | 0.003 |
| Past smoker | 2272 (38.2) | 4148 (35.6) | 6277 (36.6) | 6349 (36.6) | 0.001 |
| **Alcohol drinker – no. (%)** |  |  |  |  |  |
| Yes | 2639 (44.4) | 5361 (46.0) | 7789 (45.4) | 7908 (45.5) | 0.002 |
| No | 2836 (47.7) | 5323 (45.7) | 7911 (46.1) | 8068 (46.4) | 0.006 |
| Unknown | 472 (7.9) | 962 (8.3) | 1444 (8.4) | 1395 (8.0) | 0.014 |
| **Medication^e^ – no. (%)** |  |  |  |  |  |
| Alpha-blocker | 1090 (18.3) | 1657 (14.2) | 2856 (16.7) | 2799 (16.1) | 0.015 |
| Oral anticoagulant agent | 211 (3.6) | 390 (3.4) | 604 (3.5) | 596 (3.4) | 0.005 |
| Antiplatelet agent | 334 (5.6) | 630 (5.4) | 942 (5.5) | 974 (5.6) | 0.005 |
| Aspirin | 1638 (27.5) | 3167 (27.2) | 4884 (28.5) | 4843 (27.9) | 0.013 |
| Beta-blocker | 1329 (22.4) | 2459 (21.1) | 3842 (22.4) | 3789 (21.8) | 0.014 |
| Calcium-channel blocker | 3029 (50.9) | 5616 (48.2) | 8783 (51.2) | 8636 (49.7) | 0.030 |
| Digoxin | 64 (1.1) | 114 (0.9) | 181 (1.1) | 164 (1.0) | 0.011 |
| Diuretics | 2470 (41.5) | 4252 (36.5) | 6895 (40.2) | 6777 (39.0) | 0.025 |
| Diabetic treatment | 2486 (41.8) | 4992 (42.9) | 7414 (43.2) | 7461 (43.0) | 0.006 |
| Nitrates | 314 (5.3) | 589 (5.1) | 869 (5.1) | 867 (5.0) | 0.004 |
| Statins | 2700 (45.4) | 5350 (45.9) | 8086 (47.2) | 8067 (46.4) | 0.015 |
| **Time-related variables – mean (SD)** |  |  |  |  |  |
| Time since trial-eligible period | 430.7 (949.1) | 280.8 (764.7) | 368.9 (891.5) | 346.0 (864.9) | 0.027 |
| Number of prior ARB eligible periods | 1.2 (2.3) | 0.3 (1.0) | 0.7 (1.9) | 0.6 (1.6) | 0.023 |
| Number of prior ACEi eligible periods | 1.3 (2.4) | 1.4 (2.3) | 1.4 (2.5) | 1.4 (2.4) | 0.090 |
| Calendar year | 2011 (5.3) | 2011 (5.3) | 2011 (5.4) | 2011 (5.3) | 0.002 |
| **Healthcare utilisation^f^ – mean (SD)** |  |  |  |  |  |
| Number of GP appointments | 7.5 (27.5) | 12.4 (34.6) | 10.4 (32.0) | 10.7 (32.5) | 0.010 |
| Number of hospital admissions | 3.0 (12.6) | 2.8 (11.2) | 3.0 (12.4) | 3.1 (11.9) | 0.010 |
| **Index of multiple deprivation – no. (%)** |  |  |  |  |  |
| 1 (least) | 150 (2.5) | 314 (2.7) | 433 (2.5) | 444 (2.6) | 0.002 |
| 2 | 370 (6.2) | 675 (5.8) | 989 (5.8) | 1011 (5.8) | 0.002 |
| 3 | 924 (15.5) | 1735 (14.9) | 2609 (15.2) | 2649 (15.3) | 0.001 |
| 4 | 2131 (35.8) | 4250 (36.5) | 6228 (36.3) | 6317 (36.4) | 0.001 |
| 5 (most) | 2372 (39.9) | 4672 (40.1) | 6885 (40.2) | 6950 (40.0) | 0.003 |
| N= number of patients; no. (%)=number (percent); SD= standard deviation; SMD=standardised mean difference; BP= blood pressure; CAD=coronary artery disease; PAD=peripheral artery disease  Post-weighting N displays weighted distribution of number of patients in the two exposure groups.  Propensity-score weights are unstabilized inverse probability weights.  One third of ONTARGET participants received both ramipril plus telmisartan.  ^a^ Includes diagnosis of: MI at least 2 days prior, angina at least 30 days prior, angioplasty at least 30 days prior, CABG at least 4 years prior  ^b^ Includes diagnosis of: stroke/TIA  ^c^ Includes diagnosis of: limb bypass surgery, limb/foot amputation, intermittent claudication  ^d^ Includes DM with: retinopathy, neuropathy, chronic kidney disease, proteinuria or other complication  ^e^ Within 3 months prior to eligible start date. Antiplatelet agent= clopidogrel/ticlopidine  ^f^ Within 6 months prior to eligible start date. | | | | | |
