## Supplementary Table S5 for "Comparative effectiveness of ARB and ACEi for cardiovascular outcomes and risk of angioedema among different ethnic groups in England: an analysis in the UK Clinical Practice Research Datalink with emulation of a reference trial (ONTARGET)"

| **Supplementary Table S5** Baseline characteristics and standardised differences of trial-eligible South Asian patients after applying trial criteria included in propensity-score—weighted analysis before and after weighting for extending analysis to underrepresented groups | | | | | |
| --- | --- | --- | --- | --- | --- |
| **Characteristic** | **South Asian ethnic group** | | | | |
|  | **Before weighting** | | **After weighting** | | |
|  | **ARB**  N=10,668 | **ACEi**  N=20,137 | **ARB**  N=30,176 | **ACEi**  N=30,309 | **SMD** |
| **Age (year) – mean (SD)** | 67.7 (845) | 67.1 (8.5) | 67.4 (8.5) | 67.4 (8.5) | 0.006 |
| **Systolic BP (mmHg) – mean (SD)** | 140.4 (19.1) | 141.0 (19.3) | 141.2 (19.4) | 141.1 (19.3) | 0.008 |
| **Diastolic BP (mmHg) – mean (SD)** | 78.0 (10.7) | 78.7 (10.8) | 78.6 (10.8) | 78.6 (10.8) | 0.004 |
| **Body-mass index – mean (SD)** | 28.1 (5.0) | 27.6 (5.0) | 27.9 (4.9) | 27.8 (5.1) | 0.018 |
| **Creatinine (μmol/l) – mean (SD)** | 91.3 (31.2) | 88.0 (28.2) | 90.2 (29.4) | 90.2 (30.0) | 0.001 |
| **Female sex – no. (%)** | 5561 (52.1) | 9362 (46.5) | 14662 (48.6) | 14678 (48.4) | 0.003 |
| **Clinical history – no. (%)** |  |  |  |  |  |
| CAD^a^ | 7132 (66.9) | 13740 (68.2) | 20362 (67.5) | 20557 (67.8) | 0.007 |
| Cerebrovascular disease^b^ | 1005 (9.4) | 1986 (9.9) | 2873 (9.5) | 2907 (9.6) | 0.002 |
| PAD^c^ | 982 (9.2) | 1828 (9.1) | 2751 (9.1) | 2755 (9.1) | 0.001 |
| Diabetes | 8446 (79.2) | 15767 (78.3) | 23771 (78.8) | 23812 (78.6) | 0.005 |
| High-risk diabetes^d^ | 6068 (56.9) | 10877 (54.0) | 16878 (55.9) | 16741 (55.2) | 0.014 |
| **Smoking status – no. (%)** |  |  |  |  |  |
| Non-smoker | 4828 (45.3) | 8806 (43.7) | 13312 (44.1) | 13318 (43.9) | 0.004 |
| Current smoker | 2193 (20.6) | 4648 (23.1) | 6753 (22.4) | 6821 (22.5) | 0.003 |
| Past smoker | 3647 (34.2) | 6683 (33.2) | 10111 (33.5) | 10170 (33.6) | 0.001 |
| **Alcohol drinker – no. (%)** |  |  |  |  |  |
| Yes | 3251 (30.5) | 6273 (31.2) | 9357 (31.0) | 9375 (30.9) | 0.002 |
| No | 6488 (60.8) | 12033 (59.8) | 17776 (58.9) | 18259 (60.2) | 0.027 |
| Unknown | 929 (8.7) | 1831 (9.1) | 2823 (9.4) | 2675 (8.8) | 0.018 |
| **Medication^e^ – no. (%)** |  |  |  |  |  |
| Alpha-blocker | 1263 (11.8) | 1860 (9.2) | 3203 (10.6) | 3117 (10.3) | 0.011 |
| Oral anticoagulant agent | 337 (3.2) | 553 (2.8) | 912 (3.0) | 875 (2.9) | 0.008 |
| Antiplatelet agent | 1062 (10.0) | 1940 (9.6) | 3049 (10.1) | 3017 (10.0) | 0.005 |
| Aspirin | 3902 (36.6) | 7546 (37.5) | 11425 (37.9) | 11461 (37.8) | 0.001 |
| Beta-blocker | 2986 (28.0) | 5486 (27.2) | 8563 (28.4) | 8436 (27.8) | 0.012 |
| Calcium-channel blocker | 3757 (35.2) | 6430 (31.9) | 10408 (34.5) | 10195 (33.6) | 0.018 |
| Digoxin | 115 (1.1) | 230 (1.1) | 344 (1.1) | 351 (1.2) | 0.002 |
| Diuretics | 3395 (31.8) | 5319 (26.4) | 9061 (30.0) | 8744 (28.9) | 0.026 |
| Diabetic treatment | 5188 (48.6) | 9749 (48.4) | 14766 (48.9) | 14750 (48.7) | 0.005 |
| Nitrates | 1131 (10.6) | 2154 (10.7) | 3302 (10.9) | 3281 (10.8) | 0.004 |
| Statins | 6489 (60.8) | 12055 (59.9) | 18415 (61.0) | 18355 (60.6) | 0.010 |
| **Time-related variables – mean (SD)** |  |  |  |  |  |
| Time since trial-eligible period | 429.3 (954.2) | 274.8 (760.9) | 355.9 (884.3) | 337.6 (856.5) | 0.021 |
| Number of prior ARB eligible periods | 1.0 (1.6) | 0.2 (0.8) | 0.6 (1.2) | 0.5 (1.3) | 0.088 |
| Number of prior ACEi eligible periods | 1.2 (1.9) | 1.2 (2.0) | 1.3 (2.0) | 1.3 (2.1) | 0.001 |
| Calendar year | 2012 (5.2) | 2011 (5.2) | 2011 (5.3) | 2011 (5.1) | 0.001 |
| **Healthcare utilisation^f^ – mean (SD)** |  |  |  |  |  |
| Number of GP appointments | 7.1 (27.4) | 10.7 (32.7) | 9.6 (31.9) | 9.5 (31.2) | 0.003 |
| Number of hospital admissions | 3.1 (12.0) | 3.0 (11.8) | 3.2 (12.3) | 3.2 (12.3) | 0.004 |
| **Index of multiple deprivation – no. (%)** |  |  |  |  |  |
| 1 (least) | 1149 (10.8) | 2004 (10.0) | 3047 (10.1) | 3088 (10.2) | 0.003 |
| 2 | 1603 (15.0) | 2708 (13.5) | 4223 (14.0) | 4239 (14.0) | 0.000 |
| 3 | 2323 (21.8) | 4166 (20.7) | 6350 (21.0) | 6440 (21.3) | 0.005 |
| 4 | 2957 (27.7) | 5802 (28.8) | 8516 (28.2) | 8575 (28.3) | 0.002 |
| 5 (most) | 2636 (24.7) | 5457 (27.1) | 8040 (26.6) | 7966 (26.3) | 0.008 |
| N= number of patients; no. (%)=number (percent); SD= standard deviation; SMD=standardised mean difference; BP= blood pressure; CAD=coronary artery disease; PAD=peripheral artery disease  Post-weighting N displays weighted distribution of number of patients in the two exposure groups.  Propensity-score weights are unstabilized inverse probability weights.  One third of ONTARGET participants received both ramipril plus telmisartan.  ^a^ Includes diagnosis of: MI at least 2 days prior, angina at least 30 days prior, angioplasty at least 30 days prior, CABG at least 4 years prior  ^b^ Includes diagnosis of: stroke/TIA  ^c^ Includes diagnosis of: limb bypass surgery, limb/foot amputation, intermittent claudication  ^d^ Includes DM with: retinopathy, neuropathy, chronic kidney disease, proteinuria or other complication  ^e^ Within 3 months prior to eligible start date. Antiplatelet agent= clopidogrel/ticlopidine  ^f^ Within 6 months prior to eligible start date. | | | | | |
