## Supplementary Table S6 for "Comparative effectiveness of ARB and ACEi for cardiovascular outcomes and risk of angioedema among different ethnic groups in England: an analysis in the UK Clinical Practice Research Datalink with emulation of a reference trial (ONTARGET)"

| **Supplementary Table S6** Baseline characteristics and standardised differences of trial-eligible White patients after applying trial criteria included in propensity-score—weighted analysis before and after weighting for extending analysis to underrepresented groups | | | | | |
| --- | --- | --- | --- | --- | --- |
| **Characteristic** | **White ethnic group** | | | | |
|  | **Before weighting** | | **After weighting** | | |
|  | **ARB**  N=135,192 | **ACEi**  N=389,431 | **ARB**  N=507,289 | **ACEi**  N=515,006 | **SMD** |
| **Age (year) – mean (SD)** | 71.7 (9.1) | 71.1 (9.4) | 71.4 (9.3) | 71.3 (9.3) | 0.008 |
| **Systolic BP (mmHg) – mean (SD)** | 144.2 (20.3) | 143.8 (20.3) | 144.9 (20.6) | 144.2 (20.4) | 0.033 |
| **Diastolic BP (mmHg) – mean (SD)** | 78.8 (10.9) | 79.1 (11.0) | 79.3 (11.1) | 79.1 (11.0) | 0.016 |
| **Body-mass index – mean (SD)** | 29.2 (5.8) | 28.8 (5.8) | 29.0 (5.8) | 28.9 (5.9) | 0.010 |
| **Creatinine (μmol/l) – mean (SD)** | 94.1 (29.2) | 93.0 (207.2) | 93.9 (28.4) | 93.5 (27.9) | 0.013 |
| **Female sex – no. (%)** | 73849 (54.6) | 187167 (48.1) | 255003 (50.3) | 256177 (49.7) | 0.011 |
| **Clinical history – no. (%)** |  |  |  |  |  |
| CAD^a^ | 94954 (70.2) | 277145 (71.2) | 357856 (70.5) | 364655 (70.8) | 0.006 |
| Cerebrovascular disease^b^ | 14480 (10.7) | 41666 (10.7) | 53996 (10.6) | 54866 (10.7) | 0.000 |
| PAD^c^ | 12625 (9.3) | 36101 (9.3) | 46869 (9.2) | 47714 (9.3) | 0.001 |
| Diabetes | 80640 (59.7) | 225149 (57.8) | 296114 (58.4) | 299909 (58.2) | 0.003 |
| High-risk diabetes^d^ | 66692 (49.3) | 179831 (46.2) | 242995 (47.9) | 243302 (47.2) | 0.013 |
| **Smoking status – no. (%)** |  |  |  |  |  |
| Non-smoker | 36820 (27.2) | 100093 (25.7) | 132616 (26.1) | 134574 (26.1) | 0.000 |
| Current smoker | 31732 (23.5) | 104641 (26.9) | 130463 (25.7) | 133821 (26.0) | 0.009 |
| Past smoker | 66640 (49.3) | 184697 (47.4) | 244211 (48.1) | 246610 (47.9) | 0.005 |
| **Alcohol drinker – no. (%)** |  |  |  |  |  |
| Yes | 86807 (64.2) | 249587 (64.1) | 326979 (64.5) | 328355 (63.8) | 0.015 |
| No | 35942 (26.6) | 102414 (26.3) | 131144 (25.9) | 137854 (26.8) | 0.021 |
| Unknown | 12443 (9.2) | 37430 (9.6) | 49167 (9.7) | 48797 (9.5) | 0.007 |
| **Medication^e^ – no. (%)** |  |  |  |  |  |
| Alpha-blocker | 14736 (10.9) | 34649 (8.9) | 50488 (10.0) | 49073 (9.5) | 0.014 |
| Oral anticoagulant agent | 12507 (9.3) | 34043 (8.7) | 45941 (9.1) | 45937 (8.9) | 0.005 |
| Antiplatelet agent | 11969 (8.9) | 38410 (9.9) | 49916 (9.8) | 49874 (9.7) | 0.005 |
| Aspirin | 44853 (33.2) | 138124 (35.5) | 179775 (35.4) | 180427 (35.0) | 0.008 |
| Beta-blocker | 43419 (32.1) | 127884 (32.8) | 169400 (33.4) | 169205 (32.9) | 0.011 |
| Calcium-channel blocker | 45749 (33.8) | 123869 (31.8) | 170311 (33.6) | 168053 (32.6) | 0.020 |
| Digoxin | 5251 (3.9) | 16636 (4.3) | 20162 (4.0) | 22133 (4.3) | 0.016 |
| Diuretics | 58084 (43.0) | 153784 (39.5) | 212419 (41.9) | 209807 (40.7) | 0.023 |
| Diabetic treatment | 30647 (22.7) | 86906 (22.3) | 115932 (22.9) | 116039 (22.5) | 0.008 |
| Nitrates | 12657 (9.4) | 41176 (10.6) | 53318 (10.5) | 53411 (10.4) | 0.005 |
| Statins | 70585 (52.2) | 208064 (53.4) | 270123 (53.3) | 272453 (53.1) | 0.003 |
| **Time-related variables – mean (SD)** |  |  |  |  |  |
| Time since trial-eligible period | 443.3 (1007.9) | 269.3 (760.7) | 354.3 (907.9) | 310.1 (827.9) | 0.049 |
| Number of prior ARB eligible periods | 0.8 (1.2) | 0.1 (0.5) | 0.3 (0.8) | 0.3 (0.9) | 0.000 |
| Number of prior ACEi eligible periods | 1.4 (2.5) | 1.5 (2.4) | 1.5 (2.5) | 1.5 (2.5) | 0.073 |
| Calendar year | 2010 (5.4) | 2010 (5.3) | 2010 (5.4) | 2010 (5.3) | 0.007 |
| **Healthcare utilisation^f^ – mean (SD)** |  |  |  |  |  |
| Number of GP appointments | 5.6 (22.6) | 9.7 (28.0) | 8.3 (27.8) | 8.8 (26.7) | 0.020 |
| Number of hospital admissions | 2.9 (10.5) | 3.1 (10.3) | 3.3 (11.0) | 3.2 (10.7) | 0.014 |
| **Index of multiple deprivation – no. (%)** |  |  |  |  |  |
| 1 (least) | 29459 (21.8) | 78959 (20.3) | 104409 (20.6) | 105995 (20.6) | 0.000 |
| 2 | 30082 (22.3) | 83526 (21.5) | 109782 (21.6) | 111506 (21.7) | 0.000 |
| 3 | 26797 (19.8) | 76997 (19.8) | 100559 (19.8) | 101842 (19.8) | 0.001 |
| 4 | 25255 (18.7) | 75741 (19.5) | 97647 (19.3) | 99214 (19.3) | 0.000 |
| 5 (most) | 23599 (17.5) | 74208 (19.1) | 94892 (18.7) | 96448 (18.7) | 0.001 |
| N= number of patients; no. (%)=number (percent); SD= standard deviation; SMD=standardised mean difference; BP= blood pressure; CAD=coronary artery disease; PAD=peripheral artery disease  Post-weighting N displays weighted distribution of number of patients in the two exposure groups.  Propensity-score weights are unstabilized inverse probability weights.  One third of ONTARGET participants received both ramipril plus telmisartan.  ^a^ Includes diagnosis of: MI at least 2 days prior, angina at least 30 days prior, angioplasty at least 30 days prior, CABG at least 4 years prior  ^b^ Includes diagnosis of: stroke/TIA  ^c^ Includes diagnosis of: limb bypass surgery, limb/foot amputation, intermittent claudication  ^d^ Includes DM with: retinopathy, neuropathy, chronic kidney disease, proteinuria or other complication  ^e^ Within 3 months prior to eligible start date. Antiplatelet agent= clopidogrel/ticlopidine  ^f^ Within 6 months prior to eligible start date. | | | | | |
