## Supplementary Table S7 for "Comparative effectiveness of ARB and ACEi for cardiovascular outcomes and risk of angioedema among different ethnic groups in England: an analysis in the UK Clinical Practice Research Datalink with emulation of a reference trial (ONTARGET)"

| **Supplementary Table S7** Number of events for the primary outcome, its components, main secondary outcome and death from any cause for ARB vs ACEi using a propensity-score—weighted analysis of trial-eligible patients in CPRD Aurum after emulation and benchmarking findings against the reference trial (ONTARGET) | | | | |
| --- | --- | --- | --- | --- |
| **Outcome** | **CPRD** | | | **ONTARGET** |
|  | **ARB**  (N=151,807) | **ACEi**  (N=421,214) | **ARB vs ACEi**  (N=573,021) | **Telmisartan vs ramipril**  (N=17,118) |
|  | *Number (percent)* | | *Hazard ratio (95% CI)* | |
| Primary composite | 27327 (18.0) | 80624 (19.1) | 0.96 (0.95, 0.98) | 1.01 (0.94, 1.09) |
| Main secondary outcome | 21673 (14.3) | 65908 (15.7) | 0.94 (0.92, 0.96) | 0.99 (0.91, 1.07) |
| Myocardial infarction | 9913 (6.5) | 31069 (7.4) | 0.96 (0.94, 0.99) | 1.07 (0.94, 1.22) |
| Stroke | 6870 (4.5) | 19390 (4.6) | 0.96 (0.93, 0.99) | 0.91 (0.79, 1.05) |
| Hospitalisation for heart failure | 10029 (6.6) | 26771 (6.4) | 1.03 (1.00, 1.06) | 1.12 (0.97, 1.29) |
| Death from cardiovascular causes | 9199 (6.1) | 28355 (6.7) | 0.91 (0.89, 0.94) | 1.00 (0.89, 1.12) |
| Death from non-cardiovascular causes | 14241 (9.4) | 43391 (10.3) | 0.91 (0.89, 0.93) | 0.96 (0.83, 1.10) |
| Death from any cause | 23440 (15.4) | 71746 (17.0) | 0.91 (0.90, 0.93) | 0.98 (0.90, 1.07) |
| Doubling of serum creatinine | 5289 (3.9) | 12588 (3.4) | 1.09 (1.05, 1.13) | 1.11 (0.88, 1.39) |
| Primary composite outcome: death from cardiovascular causes, myocardial infarction, stroke, or hospitalisation for heart failure.  Main secondary outcome: death from cardiovascular causes, myocardial infarction, or stroke.  ESKD: end-stage kidney disease; GFR: glomerular filtration rate.  CPRD weighted analysis includes 1 randomly selected trial-eligible period per patient. Propensity-score—weighted with robust standard errors.  Myocardial infarction and stroke include both fatal and non-fatal events.  Only outcomes studied in ONTARGET where studied for the benchmarking analysis.  ONTARGET results are from published findings. | | | | |
