## Supplementary Table S8 for "Comparative effectiveness of ARB and ACEi for cardiovascular outcomes and risk of angioedema among different ethnic groups in England: an analysis in the UK Clinical Practice Research Datalink with emulation of a reference trial (ONTARGET)"

| **Supplementary Table S8** Comparative effectiveness of ARB vs ACEi for the primary and secondary outcomes overall and by ethnicity with a test for multiplicative heterogeneity using a propensity-score—weighted analysis of trial-eligible patients in CPRD Aurum with an on-treatment approach. | | | | | |
| --- | --- | --- | --- | --- | --- |
| **Outcome** | **Overall**  (N=573,021) | **By ethnic group** | | | |
|  |  | **Black**  (N=17,593) | **South Asian**  (N=30,805) | **White**  (N=524,623) | **P value for interaction** |
|  | *Hazard ratio (95% CI)* | | | |  |
| Primary composite | 0.95 (0.94, 0.97) | 1.00 (0.91, 1.10) | 0.95 (0.89, 1.02) | 0.95 (0.94, 0.97) | 0.581 |
| Main secondary outcome | 0.93 (0.92, 0.95) | 1.01 (0.90, 1.12) | 0.96 (0.90, 1.03) | 0.93 (0.91, 0.95) | 0.236 |
| Myocardial infarction | 0.95 (0.92, 0.98) | 1.10 (0.92, 1.32) | 0.96 (0.88, 1.06) | 0.95 (0.92, 0.97) | 0.245 |
| Stroke | 0.95 (0.92, 0.98) | 0.95 (0.81, 1.11) | 0.97 (0.85, 1.10) | 0.95 (0.92, 0.98) | 0.975 |
| Hospitalisation for heart failure | 1.01 (0.99, 1.04) | 0.98 (0.85, 1.13) | 0.97 (0.87, 1.07) | 1.02 (0.99, 1.05) | 0.579 |
| Death from cardiovascular causes | 0.90 (0.88, 0.93) | 1.14 (0.97, 1.35) | 0.94 (0.83, 1.06) | 0.89 (0.87, 0.92) | 0.013 |
| Death from non-cardiovascular causes | 0.90 (0.88, 0.92) | 0.97 (0.85, 1.12) | 0.91 (0.82, 1.02) | 0.90 (0.88, 0.92) | 0.571 |
| Death from any cause | 0.90 (0.89, 0.92) | 1.04 (0.93, 1.15) | 0.92 (0.85, 1.00) | 0.90 (0.88, 0.92) | 0.026 |
| Loss of GFR or ESKD | 1.05 (1.02, 1.08) | 1.09 (0.94, 1.26) | 1.00 (0.90, 1.13) | 1.05 (1.02, 1.08) | 0.665 |
| ESKD | 0.99 (0.94, 1.05) | 1.10 (0.88, 1.37) | 0.96 (0.80, 1.14) | 0.99 (0.94, 1.05) | 0.603 |
| Doubling of serum creatinine | 1.06 (1.02, 1.10) | 1.08 (0.89, 1.29) | 1.02 (0.88, 1.76) | 1.06 (1.02, 1.10) | 0.852 |
| Angioedema | 0.62 (0.50, 0.77) | 0.36 (0.18, 0.71) | 0.60 (0.25, 1.44) | 0.66 (0.52, 0.84) | 0.254 |
| Primary composite outcome: death from cardiovascular causes, myocardial infarction, stroke, or hospitalisation for heart failure. Main secondary outcome: death from cardiovascular causes, myocardial infarction, or stroke. Loss of GFR or ESKD defined as: 50% reduction in estimated glomerular filtration ratio (eGFR), start of kidney replacement therapy (KRT) or eGFR<15ml/min/1.73m^2^. ESKD defined as: start of KRT or eGFR<15ml/min/1.73m^2^.  ESKD: end-stage kidney disease; GFR: glomerular filtration rate.  CPRD weighted analysis includes 1 randomly selected trial-eligible period per patient. Propensity-score—weighted with robust standard errors.  Myocardial infarction and stroke include both fatal and non-fatal events.  Under on-treatment analysis, patients were additionally censored at the end of an eligible period, if they switched treatment or started dual therapy. This was denoted as date of last drug and patients were censored at this date +60 days. | | | | | |
