## Supplementary Table S9 for "Comparative effectiveness of ARB and ACEi for cardiovascular outcomes and risk of angioedema among different ethnic groups in England: an analysis in the UK Clinical Practice Research Datalink with emulation of a reference trial (ONTARGET)"

| **Supplementary Table S9** Comparative effectiveness of ARB vs ACEi for the primary and secondary outcomes overall and by ethnicity using a propensity-score—weighted analysis of trial-eligible patients in CPRD Aurum with after multiple imputation of missing values. | | | | |
| --- | --- | --- | --- | --- |
| **Outcome** | **Overall**  (N=690,166) | **By ethnic group** | | |
|  |  | **Black**  (N=21,015) | **South Asian**  (N=37,240) | **White**  (N=631,911) |
|  | *Hazard ratio (95% CI)* | | | |
| Primary composite | 0.95 (0.94, 0.97) | 1.02 (0.94, 1.11) | 0.99 (0.93, 1.04) | 0.95 (0.93, 0.96) |
| Main secondary outcome | 0.93 (0.91, 0.94) | 1.01 (0.92, 1.12) | 1.00 (0.94, 1.06) | 0.92 (0.91, 0.94) |
| Myocardial infarction | 0.95 (0.92, 0.97) | 1.08 (0.92, 1.27) | 1.00 (0.91, 1.09) | 0.94 (0.92, 0.97) |
| Stroke | 0.95 (0.93, 0.98) | 1.02 (0.88, 1.17) | 0.98 (0.87, 1.10) | 0.95 (0.92, 0.98) |
| Hospitalisation for heart failure | 1.03 (1.01, 1.06) | 1.01 (0.89, 1.15) | 1.00 (0.91, 1.10) | 1.03 (1.01, 1.06) |
| Death from cardiovascular causes | 0.90 (0.88, 0.93) | 1.13 (0.97, 1.31) | 0.99 (0.89, 1.11) | 0.89 (0.87, 0.92) |
| Death from non-cardiovascular causes | 0.91 (0.89, 0.93) | 0.93 (0.82, 1.06) | 0.94 (0.85, 1.03) | 0.90 (0.89, 0.92) |
| Death from any cause | 0.91 (0.89, 0.92) | 1.01 (0.92, 1.11) | 0.96 (0.90, 1.03) | 0.90 (0.89, 0.92) |
| Angioedema | 0.62 (0.50, 0.75) | 0.37 (0.19, 0.72) | 0.73 (0.34, 1.57) | 0.65 (0.52, 0.81) |
| Primary composite outcome: death from cardiovascular causes, myocardial infarction, stroke, or hospitalisation for heart failure. Main secondary outcome: death from cardiovascular causes, myocardial infarction, or stroke. Loss of GFR or ESKD defined as: 50% reduction in estimated glomerular filtration ratio (eGFR), start of kidney replacement therapy (KRT) or eGFR<15ml/min/1.73m^2^. ESKD defined as: start of KRT or eGFR<15ml/min/1.73m^2^.  ESKD: end-stage kidney disease; GFR: glomerular filtration rate.  CPRD weighted analysis includes 1 randomly selected trial-eligible period per patient. Propensity-score—weighted with robust standard errors.  Myocardial infarction and stroke include both fatal and non-fatal events.  Multiple imputation of chained equations for missing systolic and diastolic blood pressure and serum creatinine at baseline. | | | | |
